# Wastewater sequence data reveal national and cross-border virus transmission dynamics

**DOI:** 10.64898/2026.11.13.26368022

**Authors:** Gergely Ódor, Lukas Endler, Fabian Amman, Andreas Bergthaler

**Affiliations:** Institute for Hygiene and Applied Immunology, Center of Pathophysiology, Infectiology and Immunology, Medical University of Vienna, Austria; HUN-REN Alfréd Rényi Institute of Mathematics, Budapest, Hungary; Ludwig Boltzmann Institute for Science Outreach and Pandemic Preparedness, Medical University of Vienna, Austria; Research Center for Molecular Medicine of the Austrian Academy of Sciences, Vienna, Austria

## Abstract

Spatial epidemic models estimate disease transmission counts between populations using human mobility data, but location-to-location transmission is rarely observed directly. To address this gap, we develop mutation-counting methods to infer transmission structure from viral genetic variation measured in wastewater. In epidemic simulations with known transmission networks, our primary estimator robustly recovers spatial structure under realistic mutation-detection limits. Applied to spatiotemporal SARS-CoV-2 wastewater data from Austria, the method reveals strong seasonal reorganization, with inferred transmission becoming more geographically localized in winter and more dispersed in summer. We further identify locations with systematically higher or lower connectivity than expected from commuting, with temporal changes coinciding with regional restrictions, reopening and tourism. We then test whether such inference remains possible across national borders, recovering coherent spatial structure and temporal variation in connectivity between Austria, Switzerland and Liechtenstein. These results establish wastewater sequence data as a promising complement to mobility data for monitoring spatial transmission and detecting changes in epidemic spread associated with seasonality and interventions.

## Introduction

Network epidemic models provide a natural framework for representing the spatial spread of infectious diseases, with mobility, commuting and contact data commonly used to define connections between locations [1, 2, 3, 4]. However, such data capture opportunities for movement and interaction rather than the transmission events realized during an epidemic. The network of realized transmissions may therefore differ substantially from that implied by mobility and contact data, as transmission also depends on the nature and intensity of interactions, the infection and immunity status of individuals, and stochastic infection events [5]. Since location-to-location transmission is rarely observed directly, these differences remain difficult to quantify empirically [6]. Here, we show how wastewater sequence data can be used to estimate spatial transmission structure from the genetic variation of a pathogen.

A growing body of research has used genomic data to estimate spatial transmission patterns. Phylodynamic approaches reconstruct phylogenetic relationships between sampled genomes and use geographic annotations to infer migration or transmission between populations [7, 8, 9]. More recently, direct methods have been developed to infer transmission patterns without phylogenetic reconstruction. Spatial transmission between populations has been inferred from identical or closely related clinical consensus sequences [10, 11], while machine-learning approaches have been used to identify the spatial scales of interactions associated with viral genetic structure [12]. At the individual level, clinical sequences have been combined with observed or modelled contact-network information to reconstruct transmission structure [13, 14]. Genetic variation has also been used to identify superspreading events and infer subsequent changes in epidemic behaviour that can alter spatial transmission patterns [15]. However, these approaches generally rely on individual-level genomes obtained through clinical surveillance, which can introduce sampling biases arising from factors such as socioeconomic inequalities in testing uptake and geographic disparities in sequencing capacity [16, 17].

Wastewater surveillance offers a complementary population-level approach that can mitigate sampling biases associated with clinical testing by providing broad coverage at relatively low per-capita costs [18, 19].

During the COVID-19 pandemic, wastewater monitoring expanded rapidly, with national and regional programs established in many countries to monitor SARS-CoV-2 circulation at the population level [20]. While many surveillance programs initially focused on measuring viral concentrations to track community prevalence, wastewater sequencing was also integrated into several programs to monitor SARS-CoV-2 lineage dynamics over time and across locations [19, 21, 22, 23, 24]. In addition to monitoring circulating lineages, wastewater sequencing can capture viral genetic diversity from multiple infections within a catchment, including those missed by clinical surveillance [19, 22]. This diversity can also provide information about local epidemic activity beyond viral concentration measurements, as demonstrated by its association with community COVID-19 incidence [25].

At the same time, wastewater sequence data pose several challenges [26]. Each sample contains genetic material shed by many infected individuals, making it difficult to determine which mutations occur together on the same viral genome. Low-frequency observations may be affected by amplification, sequencing errors, and read depth, while sampling frequency and quality may vary across locations and time. Methods developed for complete individual-level genomes therefore cannot be applied directly to wastewater sequencing data without additional genome reconstruction steps [27, 28], which introduce considerable uncertainty and computational complexity.

A simpler approach to analyzing wastewater sequence data is to compare mutation profiles without reconstructing linkage between mutations. Related mutation-based approaches have previously been used in clinical genomic epidemiology to estimate epidemiological parameters [29] and infer transmission routes [30]. More recently, mutation-based approaches have been applied to wastewater sequencing data to characterize genetic diversity and spatiotemporal patterns of viral variation [31, 32, 33]. Although these studies demon-strate that mutation profiles capture meaningful temporal and geographic patterns, they do not directly evaluate how accurately mutation-based similarity recovers underlying transmission structure. Furthermore, while genetic similarity has been shown to correlate with geographic proximity [33], its relationship to independent measures of epidemiological connectivity, such as human mobility data, remains underexplored.

To address this gap, we develop interpretable mutation-counting methods that translate spatiotemporal patterns of wastewater mutations into *estimated transmission matrices* (ETMs, see Fig. 1). We establish using epidemic simulations that these estimated matrices recover spatial transmission structure under realistic observation limits. We then apply the approach to wastewater surveillance data from Austria, Switzerland and Liechtenstein, revealing seasonal reorganization of spatial transmission patterns in agreement with expectations from mobility data. By focusing on the differences between the estimated matrices and mobility, we detect location-specific deviations coinciding with regional interventions, and temporal variation in cross-border connectivity. Through these results, we demonstrate the untapped potential of wastewater sequence data as a complementary tool for monitoring realized patterns of spatial disease transmission.

**Figure 1:**
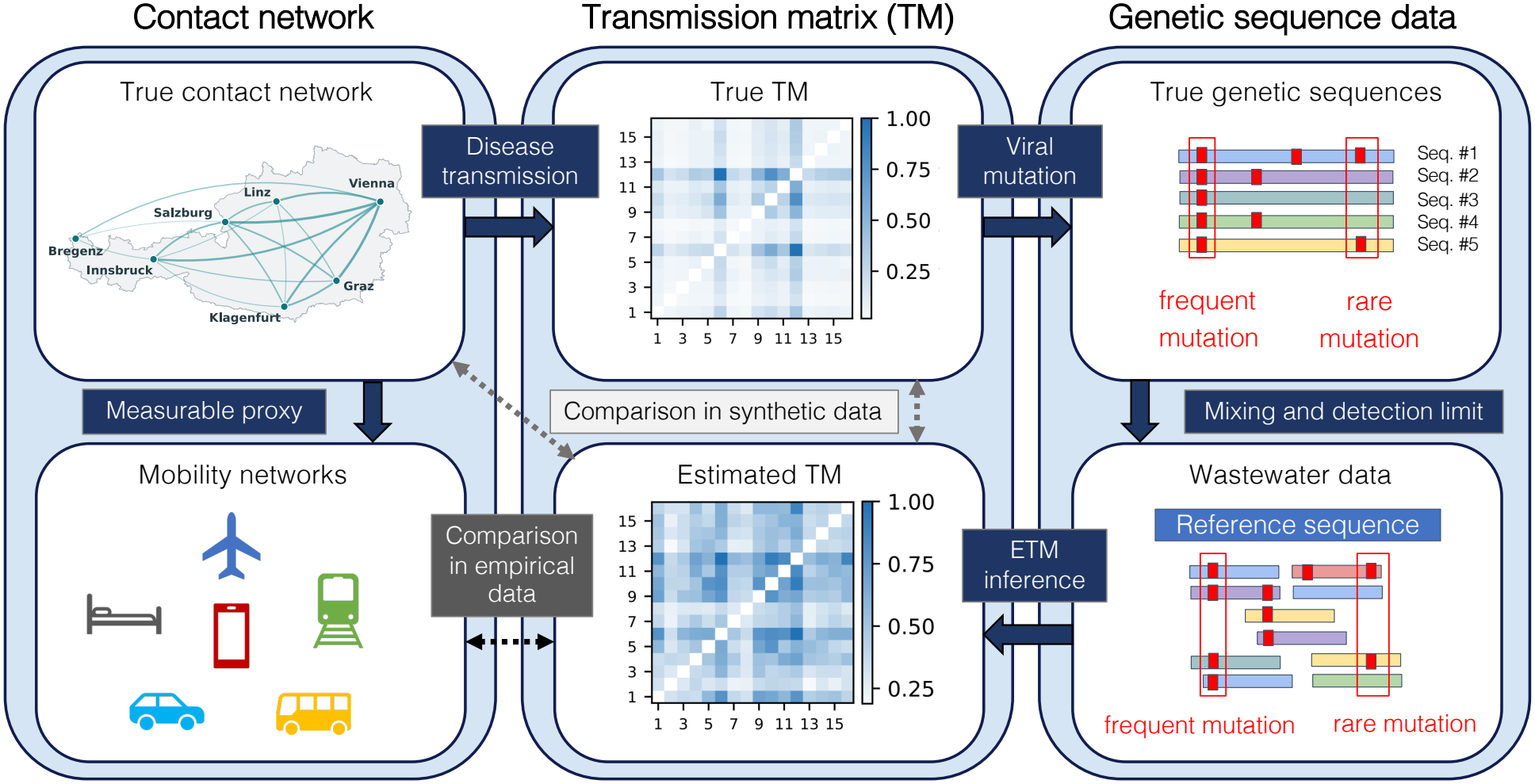
Conceptual framework for estimating and evaluating ETMs. Contact patterns between locations give rise to realized transmission through epidemic spread, which is reflected in pathogen genetic variation. Wastewater sequencing provides an incomplete observation of this variation because of within-catchment mixing, incomplete sampling and detection limits. We infer estimated transmission matrices (ETMs) from these wastewater mutation observations. In synthetic experiments, ETMs can be compared directly with the known realized transmission matrix and underlying contact structure. In empirical data, ETMs are compared with independent mobility and commuting data to identify both broad similarities in spatial transmission structure and departures from mobility-based expectations.

## Results

### Method overview

Wastewater data from Austria, Switzerland and Liechtenstein were harmonized by wastewater treatment plant (WWTP) catchment and sampling week (Methods 1.1). The Austrian dataset, which also included samples from Liechtenstein, comprised 6,534 samples collected between January 2021 and December 2024 from 48 catchments. The Swiss dataset included 1,371 samples collected between May 2021 and August 2024 from 15 catchments. We also developed an end-to-end synthetic validation pipeline to reproduce the main steps of the data generation and observation process (Fig. 1). The pipeline combined spatial contact network generation, an individual-based epidemic and mutation simulation, and a synthetic wastewater observation model with a soft allele-frequency detection limit (Methods 1.3). Both the empirical and the synthetic pipelines produced the same type of output: whether a particular substitution was observed at a given time and location, allowing the same inference methods to be evaluated in simulations and applied to empirical data.

These observations were converted into ETMs, which aim to approximate the true transmission matrix between locations, up to scaling factors. We propose two complementary methods for inferring ETMs, illustrated in Fig. 2a. We call the first approach *First-Occurrence Propagation* (FOP) counting, as it identifies mutations that are first observed in a single location and records their subsequent appearance at other locations at the next observation time as directed, one-step propagation events. FOP counting can be shown theoretically to be proportional to the true directed transmission counts under idealized transmission and observation models (Supplementary Section G). Its main limitation is that newly arising mutations may initially be carried by only a single infected individual. FOP counting therefore requires highly sensitive mutation detection and becomes less reliable when low-frequency mutations are missed.

**Figure 2:**
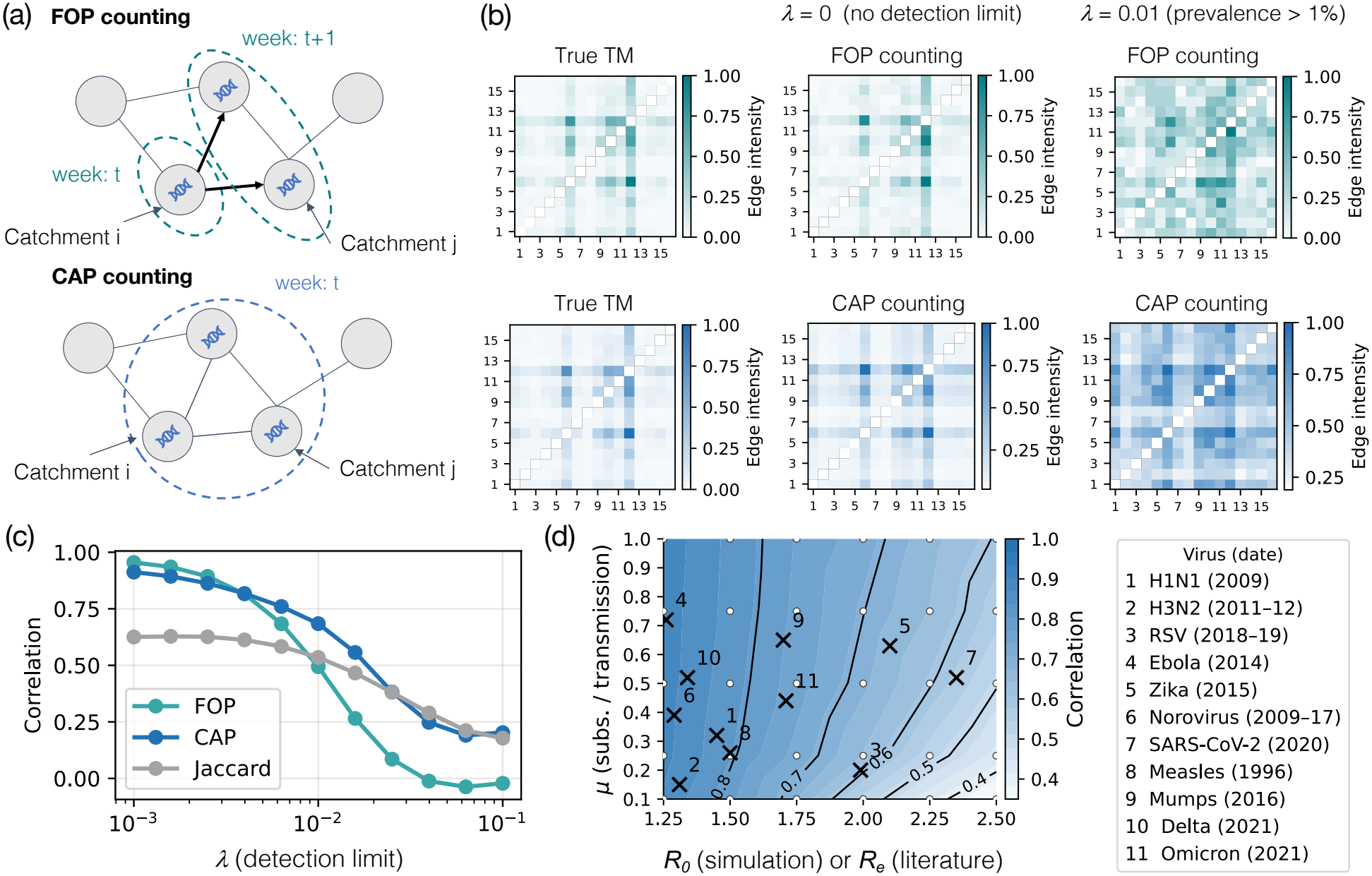
Synthetic validation of estimated transmission-matrix inference. (a) Schematic of First-Occurrence Propagation (FOP) counting, and Co-occurrence Adjusted for Prevalence (CAP) counting (Methods 1.2). (b) True transmission matrix and ETMs inferred by FOP counting (upper row) and CAP counting (lower row) from one representative simulation, shown with complete mutation observation (*λ* = 0) and a realistic allele-frequency detection limit of *λ* = 0.01. All matrices were normalized such that their maximum value is 1, and the same true transmission matrix is repeated in both rows to facilitate comparison. Within each ETM, matrix entries are interpreted relative to one another: larger values indicate stronger inferred connectivity between the 16 simulated locations, while the absolute scale is not directly interpretable. (c) Pearson correlation between the off-diagonal entries of the true transmission matrix and the inferred ETM as the detection limit *λ* is varied. Data points represent means across 500 independently seeded simulations (95% confidence intervals are too narrow to be visible). (d) CAP counting recovery across the basic reproduction number *R*_0_ and the expected number of substitutions per genome per transmission, *µ*. White circles indicate simulated parameter combinations (mean of 100 independent simulations), the colored surface shows linear interpolation between simulated values, and black contours indicate correlation levels. Numbered crosses show literature-derived reference values of the main parameters for representative viral epidemics (Methods 1.7). For these reference points, estimates of the effective reproductive number *R*_*e*_ are used instead of *R*_0_ to account for pre-existing population immunity.

We propose *Co-occurrence Adjusted for Prevalence* (CAP) counting as a more robust alternative to FOP counting. At each time point, it measures the mutations shared by each pair of locations and weights each mutation inversely by the number of locations in which it is observed. Mutations confined to a small number of locations therefore contribute more than widespread mutations, which provide little information about specific connections. CAP counting is a heuristic and does not infer direction, but it uses all informative shared mutations rather than only first-observation events. We evaluated alternative parameterizations of this prevalence weighting and compared both methods with temporally resolved Jaccard similarity as a simpler baseline (Methods 1.2) in synthetic experiments.

### Synthetic experiments

#### Recovery of spatial transmission structure

We first tested whether the inferred ETMs recovered the known transmission matrices in example simulations. While we cannot expect to recover absolute transmission counts, Fig. 2b shows that both FOP counting and CAP counting retained the broad organization of the true matrix, including at a realistic allele-frequency detection limit of *λ* = 0.01. Measuring the performance of the algorithms as the Pearson correlation between true transmission matrices and ETMs, no single approach performed best over the full range of detection limits (Fig. 2c). FOP counting excelled for *λ* values close to zero, when mutation observation was nearly complete, whereas the method based on Jaccard similarity was the best at high detection limits. CAP counting performed best at intermediate detection limits, including around *λ* = 0.01, and remained close to the best-performing approach at both extremes. Thus, although FOP counting and Jaccard were favoured under particular observation regimes, CAP counting provided the most consistently strong recovery across the range considered. This robustness, which persisted across a wide range of epidemiological and observation settings (Supplementary Fig. A.3), motivated its use in the empirical analyses.

Since recovery may also depend on pathogen and epidemic characteristics, we examined recovery across relevant combinations of the basic reproduction number *R*_0_, defined as the expected number of secondary infections generated in a fully susceptible population, and the per-transmission mutation rate *µ* (Methods 1.3.3). Fig. 2d shows that recovery was generally highest at low *R*_0_ and high *µ*. We interpret this as higher *µ* generating more mutations that can inform reconstruction of transmission structure, whereas higher *R*_0_ may reduce temporal resolution by compressing transmission events in time and leaving individual mutations at lower frequencies, making them more likely to fall below the allele-frequency detection limit (Supplementary Fig. A.2). To place this parameter landscape in biological context, we compiled literature-derived estimates of these parameters for representative viruses of public health relevance (Methods 1.7). For the epidemiological parameter, we used literature-derived estimates of the effective reproductive number *R*_*e*_ instead of *R*_0_, so that the reference points reflect transmission under the epidemiological conditions of the respective outbreaks, including population immunity and interventions. Correlations were approximately 0.8–0.9 at most reference points, suggesting that spatial transmission structure can be recovered across a broad range of plausible viral epidemic regimes.

#### Recovery of geographic localization

Since individual ETM entries can remain sensitive to incomplete and noisy mutation observations, we next evaluated whether the matrices preserved systematic properties of the underlying contact structure. We first considered spatial localization, which is commonly represented in human mobility and spatial epidemic models using gravity models that relate interactions between locations to population size and geographic distance [34, 35]. In the synthetic experiments, infection probabilities between individuals were scaled by weights *W*_*ij*_ according to the standard gravity-model formulation:

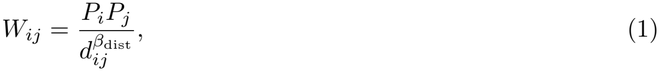

where *P*_*i*_ is the population of location *i, d*_*ij*_ is the distance between two locations, and *β*_dist_ controls how rapidly connectivity declines with distance. To estimate this distance-decay exponent, we fitted a log-linear gravity model to the positive pairwise entries *S*_*ij*_ of each inferred ETM:

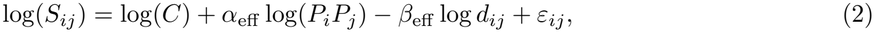

where *β*_eff_ is the fitted effective distance-decay exponent, *α*_eff_ is the population exponent, *C* is a fitted constant and *ε*_*ij*_ is a Gaussian error term. Across simulations, *β*_eff_ increased with the generating value *β*_dist_, showing that the ETMs preserved relative differences in spatial localization, although *β*_eff_ was systematically lower than *β*_dist_ (Supplementary Fig. B.1). We therefore interpret *β*_eff_ as an effective measure of spatial localization rather than as a direct estimate of the underlying distance-decay exponent. We include a robustness analysis of CAP counting to temporal variation in Supplementary Fig. B.2.

#### Detection of outlier locations

Beyond describing broad spatial and temporal structure, ETMs can also be used to identify locations whose connectivity departs systematically from what can be expected based on the general contact structure. To identify such outlying locations, we fitted an extended version of the gravity model in Eq. 2 with location-specific effects:

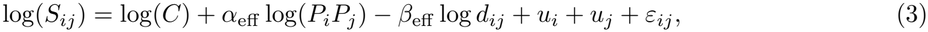

where *u*_*i*_ captures the systematic departure associated with location *i*, while *C, α*_eff_, *β*_eff_, and *ε*_*ij*_ are defined as above. For automatic detection, we included *u*_*i*_ for every site and estimated the model using the hierarchical Bayesian framework described in Methods (Eq. 15), which regularizes weakly supported deviations towards zero while allowing consistently supported departures to emerge.

We tested the location-effect model by reducing transmission involving a single site in simulations and asking whether the perturbed location could be detected. When the true transmission matrix was observed directly, detection was already close to perfect for a 20% reduction, indicating that the location-effect model itself was highly sensitive to local perturbations. Detection weakened when using ETMs inferred from simulated wastewater data, with a 50% reduction detected in approximately 50% or more of simulations, depending on the inference method and observation regime (Supplementary Fig. C.1). Interestingly, FOP counting often outperformed CAP counting for this local-detection task even for detection limits where CAP counting had higher correlation values in Fig. 2c. This indicates that CAP counting is better at preserving broad transmission structure, whereas FOP counting is more sensitive to local deviations in these simulation experiments (Supplementary Fig. A.3d).

### Empirical patterns of seasonal and location-specific transmission

#### Spatial transmission

Having validated CAP counting as the primary estimation approach, we applied it to the Austrian wastewater data. We visualized the strongest ETM connections in Fig. 3a–c (additional seasons in Supplementary Fig. F.1). The visualized networks showed clear but seasonally changing spatial organization: winter networks were dominated by geographically local links, whereas summer networks contained more long-distance connections. For comparison, we also reconstructed the networks using Jaccard similarity and FOP counting (Supplementary Fig. F.2), however, these networks were often dominated by long-distance connections that were difficult to interpret, which could be explained by their lower performance in the synthetic analyses at intermediate detection limits. We further quantified the spatial pattern inferred with CAP counting using the same gravity-model analysis as in the synthetic experiments (Eq. 2). In agreement with the network visualizations, the measured effective distance-decay exponent *β*_eff_ showed a clear seasonal signal, peaking in winter and reaching its lowest values in summer (Fig. 3d).

**Figure 3:**
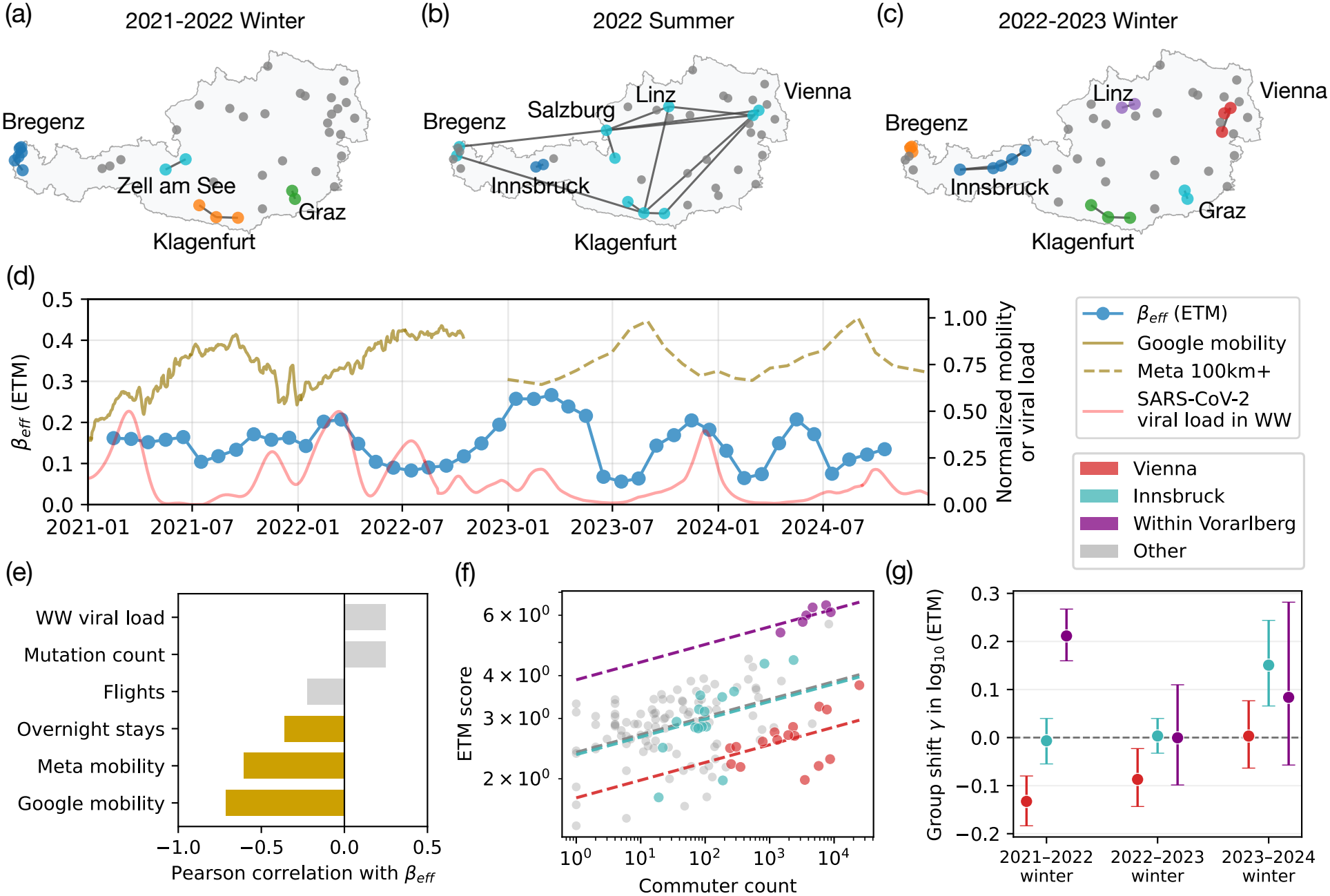
Temporal structure of Austrian ETMs and associations with human mobility. (a)– (c) Networks of the 15 strongest ETM connections among Austrian wastewater catchments in (a) winter 2021–2022, (b) summer 2022 and (c) winter 2022–2023. Node colors mark connected components of the displayed network, and smaller grey nodes denote reporting catchments not incident to one of the selected connections. Network construction is described in Methods 1.4.2; additional seasonal networks are shown in Supplementary Fig. F.1. (d) Effective distance-decay exponent *β*_eff_ obtained by fitting the gravity model in Eq. 2 to ETMs calculated in rolling three-month windows. Gold lines show normalized Google mobility (solid) and Meta 100-km-plus mobility (dashed), and the red line shows normalized SARS-CoV-2 wastewater viral load. Catchment-bootstrap confidence intervals for *β*_eff_ are shown in Supplementary Fig. F.3. (e) Pearson correlations between monthly *β*_eff_ and the indicated mobility, sequencing, and epidemiological covariates (gold bars denote *P <* 0.05; see Methods 1.6 and Supplementary Table F.1). (f) Positive ETM values versus commuter counts for catchment pairs in winter 2021–2022. Dashed lines show the group-adjusted regressions defined by Eq. 16; colors identify pairs associated with Vienna, Innsbruck, within-Vorarlberg (Austria’s westernmost federal state, covered by 6 WWTPs) connections and the remaining reference pairs. (g) Estimated group shifts *γ* after adjustment for commuter count in three consecutive winters. The dashed horizontal line denotes no group-specific shift, and error bars show 95% pair-bootstrap confidence intervals from 5,000 resamples. Corresponding fits for additional seasons and the complete rolling-window trajectories are shown in Supplementary Figs. F.5–F.6.

We compared the measured *β*_eff_ with wastewater viral load and mutation count to assess potential epidemiological and observational confounding, and with Google and Meta mobility measures to test its association with population movement. We observed that while the correlations with the potential confounders were not significant, *β*_eff_ was significantly and negatively correlated with the mobility measures (Fig. 3e). An illustrative analysis of United States clinical sequences, aggregated by state and time to emulate wastewater observations, showed a similar seasonal reorganization. For this dataset, monthly state-to-state mobility matrices were available from the SafeGraph dataset [36], and the distance-decay exponents fitted to the ETMs and the mobility networks were highly and positively correlated (*r >* 0.88, Supplementary Fig. E.1). These analyses reveal a recurring seasonal reorganization of spatial transmission: inferred connectivity became more geographically localized in winter and more spatially extended in summer. A related temporal pattern is visible in Ontario wastewater data, where the correlation between pairwise mutational similarity and geographic distance varied over time [33]. Such patterns may reflect well-established seasonal shifts in population movement and travel-driven viral introductions [37, 38]. Our analysis extends these observations by showing that seasonality was reflected in the spatial scale of the inferred transmission network itself.

#### Outlier locations

A key application of inferred transmission networks is to identify locations where epidemic connectivity departs from patterns expected from routine population movement. To identify locations with unusual connectivity, we fitted the outlier-detection model separately to each rolling window. The analysis followed the same approach as in the synthetic experiments (Eq. 15), except that connectivity was defined using commuter flows derived from Statistik Austria’s annual register-based labour market statistics [39], instead of the gravity model. The model identified several catchments whose ETM connections were consistently stronger or weaker than expected from commuting (Supplementary Fig. F.4). In 2021, the most persistent patterns involved Vienna and catchments in Vorarlberg (Austria’s westernmost federal state, bordering Switzerland and Liechtenstein), whereas Innsbruck emerged as the clearest outlying location in 2023–2024.

Based on these results, we fitted a simplified version of the location-effects model in Eq. 3, replacing the separate location-specific effects with three group-specific effects: one for connections involving Vienna, one for connections involving Innsbruck, and one for connections between catchments in Vorarlberg (Eq. 16). We fitted this model separately for each winter and found that, in winter 2021–2022, connections involving Vienna were weaker than expected from commuting, whereas those within Vorarlberg were stronger (Fig. 3f,g). However, in winter 2023–2024, connections involving Innsbruck were stronger than expected, while the Vienna and Vorarlberg effects were non-significant. We provide the complete seasonal fits and rolling-window estimates in Supplementary Figs. F.5 and F.6. The main qualitative findings remained robust when commuting data were replaced with gravity-model connectivity derived from publicly available settlement populations and geographic coordinates (see Supplementary Data and Code).

Interestingly, these observed location-specific patterns coincided with known historical events. In 2021, Vienna maintained stricter COVID-19 controls than the rest of Austria, whereas Vorarlberg served as a pilot region for the earlier reopening of gastronomy, culture and sport [40, 41, 42]. By winter 2023–2024, tourism in the Innsbruck region had strengthened again, with arrivals and overnight stays increasing by 12.2% and 6.1%, respectively, over the previous winter [43]. Although these temporal associations do not establish a direct causal link, the time-varying ETM connectivity was consistent with documented changes in restrictions, reopening and seasonal travel.

### Cross-border connectivity

We next extended the ETM analysis to wastewater data from Switzerland and Liechtenstein to examine whether transmission structure could also be inferred across national surveillance systems. Combining independently generated datasets is a more challenging setting than the within-country analyses because differences in sampling density, laboratory procedures and sequencing protocols may introduce systematic differences in the resulting ETM values. To assess whether such cross-dataset misalignments were present, we first analysed the connectivity of the main wastewater treatment plant in Liechtenstein, a smaller country geographically situated between Austria and Switzerland. Liechtenstein has strong commuting links to both countries, while its wastewater data were generated as part of the Austrian surveillance system. This provides a useful comparison, as a dataset-specific effect could make Liechtenstein appear more similar to Austrian catchments, whereas commuting patterns suggest somewhat stronger connectivity to Switzerland: in 2021, 58.9% of cross-border commuters into Liechtenstein resided in Switzerland, compared with 36.9% in Austria [44].

As in the within-Austria analyses, the combined networks retained a clear spatial organization, with more geographically localized connections in winter and more long-distance connections in summer (Fig. 4a–c). In winter 2021–2022, Liechtenstein appeared particularly strongly connected to neighbouring catchments in Vorarlberg (Fig. 4c,d), and summarizing ETM values by regional pair class showed that Liechtenstein– Vorarlberg connectivity was strongest in this first winter and decreased in subsequent years (Fig. 4e). This pattern could in principle be influenced by the denser sampling of Austrian catchments, particularly in Vorarlberg. However, repeating the comparison using a balanced subset of well-sampled Austrian and Swiss catchments preserved the stronger Liechtenstein–Austria connectivity (Supplementary Fig. F.7a), indicating that sampling density alone was not sufficient to explain the pattern. In the same balanced dataset, within-Austria, within-Switzerland and Liechtenstein–Austria ETM values generally occupied a higher range, whereas Austria–Switzerland and Liechtenstein–Switzerland comparisons were persistently lower (Fig. F.7b). This pattern is consistent with a systematic cross-dataset misalignment and suggests that absolute ETM magnitudes are not fully comparable between the independently generated Austrian/Liechtenstein and Swiss datasets.

**Figure 4:**
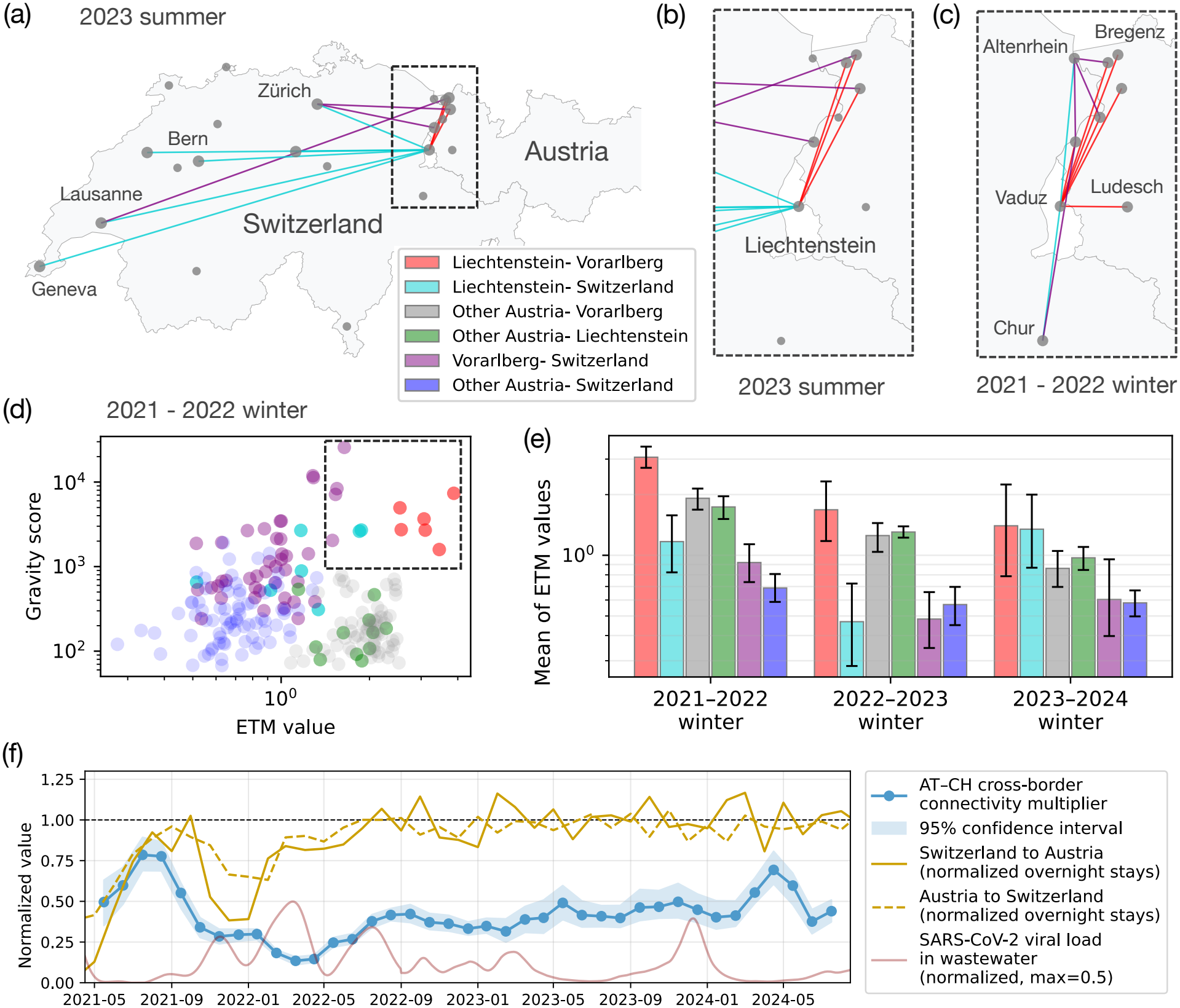
Spatio-temporal cross-border ETMs in Austria, Switzerland and Liechtenstein. (a)–(c) Spatial networks of the 12 strongest positive ETM connections among wastewater catchments in Switzerland, Vorarlberg and Liechtenstein, excluding pairs involving Austria outside of Vorarlberg. (a) Full network for summer 2023. (b) Close-up of Liechtenstein and Vorarlberg for summer 2023. (c) Close-up of the same area for winter 2021–2022. Larger nodes denote catchments incident to at least one displayed edge, whereas smaller nodes denote reporting catchments with no edge among the top 12. Edge colors indicate regional pairings (see legend). (d) ETM values versus gravity-model scores for individual catchment pairs in winter 2021–2022. Gravity scores were calculated as described in Methods 1.5. The dashed box marks the 12 connections shown in (c). (e) Geometric mean of positive ETM values for six regional pair classes across three winters. Error bars show 95% catchment-bootstrap confidence intervals from 2,000 resamples. (f) Austria–Switzerland cross-border connectivity multiplier estimated in rolling three-month windows after adjustment for population, geographic distance and country-specific effects using Eq. 17. Shading shows 95% stratified catchment-bootstrap confidence intervals from 1,000 resamples, and the dashed horizontal line at 1 marks the reference value at which cross-border connectivity matches the within-country gravity expectation. Gold lines show normalized overnight stays in both directions, and the red line shows normalized SARS-CoV-2 wastewater viral load. Only complete three-month windows are shown, and Liechtenstein was excluded from the analysis.

We therefore interpreted the broader Austria–Switzerland analysis with particular caution regarding the absolute magnitude of cross-country connectivity. We fitted a gravity model that accounted for population, geographic distance and country-specific baseline differences before estimating the additional effect associated with an Austria–Switzerland connection in rolling three-month windows (Eq. 17), yielding a *cross-border connectivity multiplier*. The estimated multiplier remained below the within-country expectation throughout the study period (Fig. 4f). However, because this term may capture both epidemiological differences across the national border and systematic differences between the two datasets, its absolute magnitude cannot be interpreted as a pure border effect. Its temporal variation may nevertheless reflect changes in cross-border connectivity under the assumption that the cross-dataset misalignment remains approximately stable over time.

The inferred cross-border connectivity multiplier showed clear temporal variation, with a pronounced reduction during winter and spring 2021–2022 (Fig. 4f). This decrease coincided with a reduction in bilateral overnight stays between Austria and Switzerland obtained from independent data sources. The same period also coincided with renewed pandemic controls, including an Austrian lockdown and stricter Swiss entry requirements [45, 46]. Although this temporal correspondence does not establish causality, it suggests that temporal changes in cross-border connectivity can remain informative despite systematic differences between the national datasets. Together with the preserved spatial structure, these results demonstrate the potential of ETMs for cross-border transmission analysis, particularly as wastewater surveillance protocols become more standardized across countries.

## Discussion

We showed that spatiotemporal mutation observations from pooled wastewater samples can be used to infer estimated transmission matrices that retain information about the spatial structure of an epidemic. To estimate these matrices we introduced two interpretable mutation-counting approaches, FOP and CAP counting, and evaluated them extensively in simulation analyses. FOP counting uses the sequential appearance of mutations to estimate directed connections and has a direct theoretical relationship to transmission under idealized conditions. CAP counting measures prevalence-adjusted mutation co-occurrence and is symmetric, but recovered the known transmission matrices more reliably under realistic observation limits. We therefore used CAP counting for the main empirical analyses to compare transmission structure inferred from genetic sequences with independent mobility data, examining both broad similarities and location-specific differences.

Several of our results point to similarities between the inferred transmission structure and independent measures of population movement. In Austria, the effective distance-decay exponent varied seasonally and was correlated with temporal Google and Meta mobility indicators. We note that while the correlation of the mobility measures and the distance-decay exponent is reassuring, their comparison is inherently indirect. A more direct comparison was possible in the United States, where spatiotemporal SafeGraph mobility matrices were available, and the distance-decay exponent of the mobility matrices could also be computed. Although this analysis used clinical sequences aggregated to resemble wastewater observations, the high correlation between the distance-decay exponents of the two datasets indicates similar temporal variation in their spatial localization. At the international scale, the Austria–Switzerland cross-border connectivity multiplier also varied similarly to bilateral overnight stays, including a marked decrease during winter and spring 2021–2022. These comparisons show that ETMs recover spatial and temporal patterns that are also present in independent mobility data.

Differences between ETMs and mobility networks may be particularly informative, as they can identify locations where routine movement does not translate into transmission as expected. Their interpretation, however, depends on the mobility dataset used for comparison. For Austria, we compared the ETMs with a static post-pandemic commuting matrix, which captures routine work-related mobility but not the temporal changes in movement and behaviour observed during the study period. The patterns involving Vienna, Vorarlberg and Innsbruck coincided with documented changes in restrictions, reopening and seasonal tourism, although these associations do not establish a causal explanation. Spatiotemporal mobility networks would allow finer comparisons and could help distinguish changes in movement from other factors affecting transmission, such as regional immunity, contact behaviour or susceptibility. However, even spatiotemporal mobility datasets have important limitations. Mobile-device and platform data may represent only selected population groups or types of movement, and even detailed movement records do not measure the specific encounters through which infections spread. ETMs and mobility networks should therefore be viewed as complementary: mobility data describe selected opportunities for movement and contact, whereas ETMs summarize the spatial structure reflected in the similarity of pathogen genetic variation.

The population-pooled nature of wastewater surveillance also creates opportunities for cross-border analyses. Wastewater sampling is less dependent than clinical surveillance on individual testing behaviour and access to healthcare, and similar measurements can be collected independently in neighbouring countries. By combining Austrian and Swiss wastewater data, we constructed a joint ETM whose temporal variation was consistent with bilateral overnight-stay data. Such analyses are nevertheless difficult because national surveillance programs can differ in sampling density, laboratory procedures, sequencing equipment and detection sensitivity. Interlaboratory comparisons have similarly shown that wastewater variant detection depends on adequate sequencing coverage and continued quality control [47]. These differences may have contributed to Liechtenstein appearing more closely connected to Austrian than to Swiss catchments in our analysis. Despite these limitations, the recovery of coherent cross-border structure from independently generated national datasets shows the potential of coordinated international wastewater surveillance.

Several additional limitations affect the interpretation of the inferred matrices. CAP counting is symmetric and cannot determine the direction of movement or transmission. Its performance also depends on dense, repeated and sufficiently comparable sequencing across locations and time. Although we assessed sensitivity to missing catchment–time reports (Supplementary Fig. A.3h), repeated introductions from outside the sampled region or a systematic lack of observations from highly connected catchments could still bias inferred connectivity. The simulations also showed that the distance-decay exponent fitted to an ETM was attenuated relative to the exponent of the generating contact matrix. The fitted *β*_eff_ should therefore be interpreted as a comparative measure of spatial localization rather than as an unbiased estimate of the underlying contact process. Furthermore, broad transmission structure was recovered more reliably than local deviations, with FOP counting sometimes outperforming CAP counting for detecting localized changes. Future estimators could improve CAP counting by incorporating some of the temporal information recovered by FOP counting. The interpretation of the empirical results has additional limitations: comparisons depend on the mobility data available, and associations with policies or travel activity remain observational. Finally, the current analyses are largely restricted to SARS-CoV-2, for which unusually extensive longitudinal sequencing data are available.

Future work should apply these methods to additional pathogens, geographic settings and intervention periods. In particular, combining harmonized wastewater sequencing with detailed spatiotemporal mobility networks could reveal new biological and public-health insights into when transmission follows standard mobility patterns and when it is altered by interventions, behavioural changes, immunity or administrative borders. Our results therefore support extending coordinated wastewater sequencing within and across countries as a new source of information on spatial epidemic spread, especially when direct observations of transmission are unavailable.

## Methods

### 1.1 Wastewater data and preprocessing

SARS-CoV-2 wastewater sequencing data from Austria and Liechtenstein were generated and processed as described in [21]. Raw wastewater sequencing data from Switzerland were generated using a different sequencing protocol [22] and subsequently processed with the same computational pipeline for mutation calling as the Austrian dataset [21]. Briefly, reads were trimmed with BBDUK and overlapping read pairs were corrected with BBMERGE from the BBTools suite (v. 38.90) [48]. Reads were mapped to a combined human GRCh38 and SARS-CoV-2 reference genome (RefSeq: NC 045512.2) using BWA-MEM (v. 0.7.17) [49]. Primer sequences were masked with iVar (v. 1.2.3) [50], and mutations were called using LoFreq (v. 2.1.2) [51], as described previously [21].

Mutations were represented by genomic position, reference allele, and alternative allele. Sample dates were assigned to weekly bins starting on Monday. We retained single-nucleotide substitutions and excluded samples with a low fraction of reads mapping to SARS-CoV-2. If multiple samples were available for the same catchment and week, the sample with the highest fraction of mapped viral reads was retained. An allele-frequency threshold of 0.01 was applied to the Austrian dataset. For the joint Austria–Switzerland analysis, the datasets were harmonized to a common set of columns and concatenated, and an allele-frequency threshold of 0.02 was applied to account for differences in sequencing protocols.

WWTP catchment coordinates and connected populations were taken from the associated metadata. Geographic distances were calculated as geodesic distances on the WGS84 ellipsoid. Further source-specific preprocessing and the processing of the United States clinical-sequence data are described in the Supplementary Materials.

### 1.2 Estimated transmission matrices

We quantified relationships between locations using two proposed matrix-based measures, FOP counting and CAP counting, and temporally resolved Jaccard similarity as a baseline. All three measures were calculated from observations (*m, t, i*) indicating that mutation *m* was observed at time *t* in location *i*. Matrices used a common set of *K* locations and had zero diagonal entries.

#### FOP counting

FOP counting records directed one-step temporal propagation. For each mutation *m*, we determined its first observed time *t*_0_(*m*) and retained it only when it was first observed in exactly one source location, *s*(*m*). At the next global observation time, *t*_1_(*m*), the mutation contributed a directed event *s*(*m*) *→ j* for every other location *j* in which it was observed. Let *c*_*ij*_(*t*) be the number of such events from source *i* to target *j* whose target time is *t*. To account for incomplete reporting, we defined *O*_*ij*_ as the set of consecutive-time observation opportunities for which source *i* was represented at the preceding global time point and target *j* was represented at time *t*. The FOP counting score was

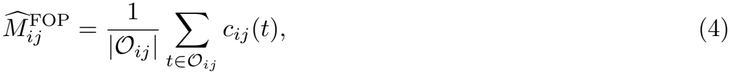

with 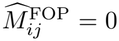 when *O*_*ij*_ was empty or no propagation event was observed. Observation opportunities with no event contributed zero. FOP counting is directed; target locations index matrix rows and source locations index columns.

#### CAP counting

CAP counting measures symmetric relatedness from shared mutations. At each time *t*, we formed a binary mutation-by-location matrix *X*(*t*), with *X*_*mi*_(*t*) = 1 when mutation *m* was observed in location *i*. We defined the prevalence of mutation *m* as *k*_*m*_(*t*) = ∑ _*i*_ *X*_*mi*_(*t*) and assigned the prevalence-adjusted weight

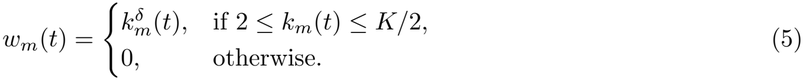

Here, *δ* controls how strongly widespread mutations are downweighted. We used *δ* = *−*1 in the main analyses, with a sensitivity analysis on *δ* included in Supplementary Fig. A.1.

The weighted overlap of locations *i* and *j* at time *t* was defined as *r*_*ij*_(*t*) = Σ_*m*_ *w*_*m*_(*t*)*X*_*mi*_(*t*)*X*_*mj*_(*t*). Let _*ij*_ denote the time points at which both locations had at least one retained mutation observation. The CAP counting score was defined as

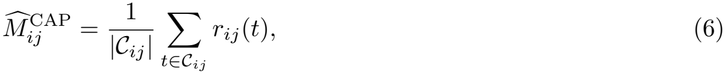

with 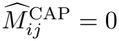 when *C*_*ij*_ was empty. Note that CAP counting is symmetric by construction.

#### Jaccard similarity

As a simpler baseline, we calculated the Jaccard similarity between the mutation sets observed in locations *i* and *j* at each time point,

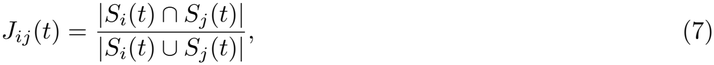

where *S*_*i*_(*t*) is the set of mutations observed in location *i* at time *t*. The Jaccard ETM entry was computed as

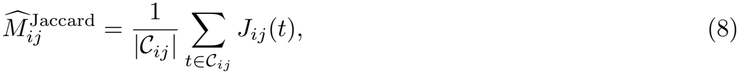

using the same co-observed time points *C*_*ij*_ as for CAP counting, with a score of zero when *C*_*ij*_ was empty.

### 1.3 Synthetic validation

#### 1.3.1 Network model

Synthetic populations contained *N* = 10,000 individuals assigned to *K* = 16 locations positioned on a 4 *×* 4 grid. Location population sizes followed a deterministic Zipf rank–size distribution, 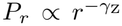, where *r* denotes the rank of a group by population size, and *γ*_Z_ is the Zipf exponent controlling the heterogeneity in group sizes. Individuals in different groups *i* ≠ *j* were connected by a complete network, with group-level interaction weights

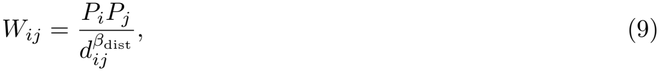

where *d*_*ij*_ denotes the distance between cities. Within-group interactions were assigned weights 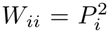 . The distances *d*_*ij*_ were defined as the Euclidean distances plus one, ensuring that *d*_*ij*_ *>* 1 for *i ≠ j*, so that the interaction weight per individual pair was higher within groups than between groups for *β*_dist_ *>* 0. By default, we chose *β*_dist_ = 1 and *γ*_Z_ = 1, with robustness analyses in Supplementary Fig. A.3c and g.

#### 1.3.2 Epidemic model

We simulated a discrete-time individual-based SIR process with transmission probabilities determined by the source and target groups. Approximately 0.5% of individuals were initialized as infected. At each update, infectious individuals at location *i* could infect susceptible individuals at location *j* with probability

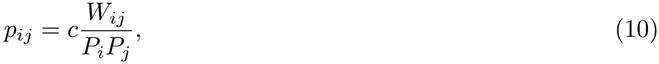

and then recovered with probability 1 at the end of the update. The scaling constant *c* was chosen so that the dominant eigenvalue of the corresponding next-generation matrix equalled the specified basic reproduction number *R*_0_. The infector, infection time and group of each infected individual were recorded as a directed infection tree (or forest). The true transmission matrix was obtained by counting recorded transmissions between source and target groups and normalizing the matrix to sum to one.

#### 1.3.3 Mutation model

Each initially infected individual was assigned the same reference sequence, and each newly infected individual inherited the sequence of its infector. Synthetic sequences contained *L* = 1,000 sites. At each transmission, the number of site updates was sampled as

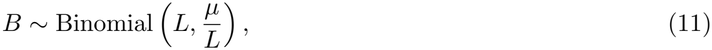

so that E[*B*] = *µ*. Selected sites were changed to one of the three alternative states. The parameter *µ* denotes the expected number of new nucleotide mutations introduced per transmission.

#### 1.3.4 Wastewater observation model

To represent short-term persistence of viral material in wastewater, each infected individual’s sequence contributed to the synthetic wastewater observation at its infection time and for one additional simulation time step. We denote this observation window by *W* = 2. Thus, at time *t*, the mutation mixture for a group was calculated from individuals infected at times *t* and *t −* 1. This persistence affected only the wastewater observation process: individuals remained infectious for a single epidemic update and could transmit only during that update. As a sensitivity analysis, we also simulated the observation model with *W* = 1, in which each sequence contributed only at the individual’s infection time in Supplementary Fig. A.3b.

Simulated sequences were aggregated by group and time point. Let 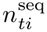 denote the total number of sequences contributing to the wastewater observation for group *i* at time *t*, and let 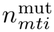 denote the number of these sequences carrying mutation *m*. The allele frequency of mutation *m* at time *t* in group *i* was then calculated as

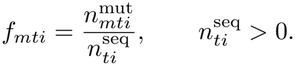

For a detection limit *λ >* 0, mutation observations were retained independently with probability

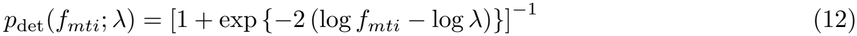

motivated by logistic detection models used in wastewater surveillance [52]. Here, *λ* corresponds to the allele frequency at which the probability of detection is 0.5. We used *λ* = 0 to denote complete observation, and *λ* = 0.01 as the default value for the detection limit, as computational benchmarks suggest that mutations at frequencies around 1% can be detected with sufficient sequencing coverage [53]. Additional perturbations removed complete group–time observations with a specified probability or introduced positions that appeared mutated in all sampled sequences. These observation mechanisms are illustrated in Supplementary Fig. A.2, with sensitivity analysis in Supplementary Fig. A.3. Unless a parameter was varied, we used *R*_0_ = 2, *µ* = 0.5, *β*_dist_ = 1 and *λ* = 0.01.

### 1.4 Evaluation and visualization

#### 1.4.1 Comparison of ETMs and true TMs in simulations

For each simulation, agreement between the true transmission matrix *M* and an inferred ETM 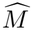 was measured by the Pearson correlation of their finite off-diagonal entries,

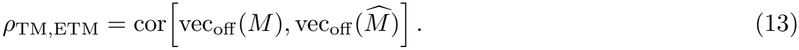

#### 1.4.2 Network visualization and rolling-window ETMs in real data

For spatial visualization, ETMs inferred by CAP counting were represented as undirected networks, and a number of the strongest positive connections (specified in each figure legend) was retained. All reporting locations were displayed; locations incident to a retained edge were drawn as larger nodes, whereas locations without a retained edge were drawn at reduced size. Unless stated otherwise in the figure legend, node colors indicate connected components of the displayed network.

For temporal comparisons, ETMs were calculated in inclusive rolling three-month windows advanced in monthly increments. To keep the same catchments across windows, we only retained catchments with at least one sample in every rolling three-month window. In Figure 3d-g, we retained 17 Austrian catchments, and in Figure 4f, we retained 17 Austrian and 5 Swiss catchments.

### 1.5 Statistical inference of spatial transmission structure

#### 1.5.1 Gravity model

For locations *i* and *j*, we modelled their positive pairwise ETM value, *S*_*ij*_, as

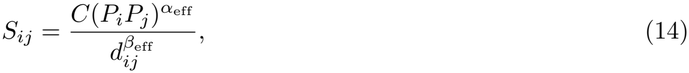

where *P*_*i*_ and *P*_*j*_ are connected populations, *d*_*ij*_ is geographic distance, *α*_eff_ is the population-scaling exponent and *β*_eff_ is the effective distance-decay exponent. The model was fitted by ordinary least squares after natural-log transformation in Eq. 2. Each unordered pair was used once, and only finite positive ETM values, populations and distances were retained. Models were fitted to the full observation period and separately to each rolling window. Rolling estimates were assigned to window midpoints. Uncertainty in rolling estimates was assessed by resampling catchments with replacement and refitting Eq. 2. For Fig. 4d, gravity scores were calculated as 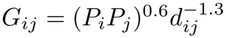.

#### 1.5.2 Bayesian location-effect model

To identify locations whose pairwise connectivity departed systematically from that expected from population movement or spatial structure, we used a hierarchical Bayesian regression model with additive location-specific effects, motivated by statistical models for pairwise network data [54]. For each analysis unit (a simulation replicate or an empirical rolling window), we modelled

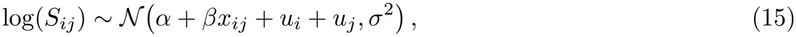

where *x*_*ij*_ describes expected pairwise connectivity, and *u*_*i*_ is the location-specific departure after accounting for this predictor. Location effects were constrained to sum to zero and assigned a hierarchical prior that induces partial pooling towards zero [55].

For the synthetic perturbation experiments, the predictor term was extended to include population and geographic distance, log(*P*_*i*_*P*_*j*_) and *−* log *d*_*ij*_. For the truly perturbed location, detection was based on the posterior probability *P* (*u*_*i*_ *<* 0 | *D*), where *D* denotes the observed data used to fit the model. The detection threshold was calibrated from unperturbed simulations to an approximately 5% false-detection rate. For the Austrian wastewater analysis, each rolling window was fitted separately using the mean-centred log-transformed commuter count as the predictor of expected connectivity. Full model specification and fitting details are provided in Supplementary Section C.

#### 1.5.3 Commuting-adjusted group effects

To quantify departures for selected Austrian locations or groups, we compared Austrian ETMs with symmetrized commuter counts using the same set of positive unordered pairs available in all three analysed winters. For each winter, we fitted

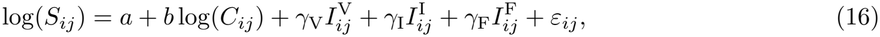

where *C*_*ij*_ is the commuter count, 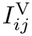 and 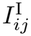 identify pairs involving Vienna and Innsbruck, respectively, and 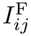 identifies pairs within Vorarlberg. All other pairs formed the reference category. The corresponding coefficients measure whether these connections were systematically higher or lower than expected from commuter flow. Confidence intervals for the group coefficients were obtained by bootstrap resampling of catchment pairs.

#### 1.5.4 Austria–Switzerland border model

For the Austria–Switzerland analysis, we augmented the gravity model as

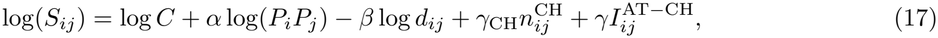

where 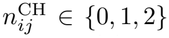 is the number of Swiss endpoints and 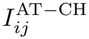 identifies Austria–Switzerland pairs.

We report exp(*γ*) as the multiplicative cross-border change relative to the fitted within-country gravity expectation. The model was fitted separately in rolling three-month windows; Liechtenstein was excluded and only complete windows were retained for Fig. 4f. Uncertainty was assessed by separately resampling Austrian and Swiss catchments with replacement and refitting Eq. 17. For the regional comparison in Fig. 4e, positive ETM values were summarized by their geometric mean within the six regional pair classes shown in the figure. Confidence intervals were obtained by resampling the contributing catchments with replacement within each class.

### 1.6 Accommodation and mobility data

The Austrian commuter matrix was aligned to wastewater-catchment names, its diagonal was set to zero and directional flows were averaged to obtain a symmetric matrix. Temporal comparisons used Google transitstation mobility [56], the Meta 100-km-plus mobility index [57], international-flight counts [58], SARS-CoV-2 wastewater viral load [59] and monthly mutation counts. The Google series was smoothed with a seven-day rolling mean before monthly aggregation; the flight series was supplied as a seven-day count. Time series were normalized only for visualization as described in the figure legends.

Monthly tourism data were obtained from Statistik Austria [60] and the Swiss Federal Statistical Office [61]. For travel from Switzerland to Austria, overnight stays were summed across Austrian federal states for visitors recorded as originating from Switzerland and Liechtenstein. For travel from Austria to Switzerland, overnight stays by Austrian residents were extracted for Switzerland as a whole. Observations through December 2024 were analysed, and each direction was normalized separately to the corresponding calendar month in 2025. Because the two national datasets do not cover identical accommodation sectors, absolute levels were not compared directly between directions. The source data and normalization are shown in Supplementary Fig. F.8. For temporal comparisons, rolling *β*_eff_ estimates were assigned to window midpoints and converted to monthly means. Mutation counts, wastewater viral load, international flights, overnight stays, Google mobility and Meta mobility were likewise aggregated to monthly resolution. Pearson correlations were calculated using pairwise-complete monthly observations, with two-sided tests used to compute *P* values. No correction for multiple comparisons was applied.

### 1.7 Biological reference values for *R*_e_ and *µ*

To place the synthetic parameter landscape in biological context, we annotated Fig. 2d with representative literature-derived reference coordinates for human viruses and viral variants. The epidemiological coordinate was based on context-specific estimates of the effective reproduction number *R*_*e*_ (or *R*_*t*_) from defined epidemics, outbreaks or seasons; where relevant, an outbreak-onset *R*_0_ was shown and identified as such in Supplementary Table D.1. Because the synthetic epidemics had no pre-existing immunity and only approximately 0.5% of individuals were infected at initialization, the specified *R*_0_ closely approximates the initial *R*_*e*_; the simulated parameter range is therefore shown on an *R*_*e*_ axis for comparison. The evolutionary coordinate *µ* denotes the expected number of substitutions per genome per transmission. Representative values were calculated as 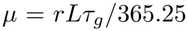, where *r* is the substitution rate per site per year, *L* is genome length and *τ*_*g*_ is the mean generation time in days, using parameter estimates compiled by Tran-Kiem and Bed-ford [62]; norovirus parameters were obtained separately (Supplementary Table D.1). The epidemiological and evolutionary coordinates were obtained independently and do not necessarily refer to the same out-break, population or time period, and should therefore be interpreted as representative biological reference coordinates rather than jointly estimated pathogen-specific parameter pairs.

### 1.8 Use of generative AI

ChatGPT (OpenAI) was used to assist with literature searches, Python code development for data processing and visualization, initial drafting of methodological descriptions, and language editing. All AI-assisted code and text were verified and revised by the authors. The final manuscript was reviewed and approved by all authors, who take full responsibility for its scientific content, analyses, interpretations, and conclusions.

## Data Availability

Data availability is described in detail in the Data Availability section of the manuscript.

https://www.ebi.ac.uk/ena/browser/view/PRJEB48985

https://www.ebi.ac.uk/ena/browser/view/PRJEB44932

https://doi.org/10.55876/gis8.261030he

https://www.google.com/covid19/mobility/

https://ai.meta.com/ai-for-good/datasets/movement-distribution-maps/

https://www.statistik.at/atlas/pendler/

https://github.com/GeoDS/COVID19USFlows-WeeklyFlows

https://www.statistik.at/en/statistics/tourism-and-transport/tourism/accommodation/arrivals-overnight-stays

https://statistik.admin.ch/en/asset/36997223

## Data Availability

All raw wastewater sequencing data used in this study are available under the ENA accession numbers: PR-JEB48985 and PRJEB44932. The identifiers and associated metadata of the 7,905 wastewater samples used in this study will be provided in Supplementary Data 1 upon acceptance of the manuscript for publication. Austrian SARS-CoV-2 wastewater viral-load data are available from the National SARS-CoV-2 Wastewater Monitoring Programme [59]. All clinical genome sequences and associated metadata are accessible through GISAID’s EpiCoV database. To view the contributors of each individual sequence with details such as accession number, Virus name, Collection date, Originating Lab and Submitting Lab and the list of Authors, visit https://doi.org/10.55876/gis8.261030he.

Austrian accommodation data are available from Statistics Austria [60], and Swiss tourism data from the Swiss Federal Statistical Office PXWeb API [61]. Austrian commuting data are available through the Statistics Austria Pendleratlas [39]. Historical mobility data are available from the Google COVID-19 Community Mobility Reports [56], and Movement Distribution Maps from Data for Good at Meta are available through the Humanitarian Data Exchange (HDX) [57]. Commercial aviation data are available through the World Bank Global Aviation Dashboard [58], and the SafeGraph mobility dataset is available at [63].

Numerical data underlying the main figures will be provided in three supplementary Excel files upon acceptance of the manuscript. Supplementary Data 2 contains simulation results and reference parameters related to Figure 2. Supplementary Data 3 contains estimated transmission matrices, settlement coordinates and populations, mobility indicators, and wastewater viral loads related to Figure 3. Supplementary Data 4 contains estimated transmission matrices, settlement coordinates and populations, wastewater viral loads, and processed tourism indicators related to Figure 4. Although the WWTP coordinates and the catchment populations used in the main analyses cannot be shared without permission from the data providers, the main qualitative findings are reproducible using the provided settlement-level data. Variable descriptions, data sources, and processing steps will be included in Supplementary Data 5.

## Code Availability

Python implementations of the FOP and CAP estimators, example simulations, and code to reproduce the main Figures 2–4 using Supplementary Data 2–4 will be made available upon acceptance of the manuscript.

## Acknowledgements

We thank Martin Bicher, Domonkos Czifra, Berna De Vries, Emily Grubb, Marton Karsai, Niki Popper, Patrick Thiran, and Julia Vierheilig for their insightful comments and remarks. The wastewater surveillance programs in Austria and Liechtenstein were conducted by the Austrian Federal Ministry of Education, Science and Research, the Austrian Federal Ministry of Social Affairs, Health, Care and Consumer Protection and the Principality of Liechtenstein. This study was supported by the Austrian Science Fund (FWF) Cluster of Excellence “Microbiomes drive planetary health” (doi: 10.55776/COE7; to A.B.) and the FWF grant P34579 (doi: 10.55776/P34579; to A.B.). A.B. and F.A. were supported by the Ludwig Boltzmann Institute for Science Outreach and Pandemic Preparedness, funded by the Ludwig Boltzmann Gesellschaft, Medical University of Vienna, Stadt Wien, and Gesundheit Österreich GmbH (GÖ G).

## Author contributions

G.O. performed the data analysis and synthetic simulations, created the visualizations, and wrote the first draft of the manuscript. L.E. and F.A. performed wastewater data preprocessing. All authors contributed to the design of the research and reviewed the final manuscript.

## Competing interests

The authors declare no competing interests.

## Notes

### Competing Interest Statement

The authors have declared no competing interest.

### Author Declarations

The Ethics Commission of the City of Vienna waived ethical approval for a preceding study using the same genomic data collection pipeline (doi: 10.1038/s41587-022-01387-y), as the study involved only non-identifiable biological material and data and therefore did not require ethics committee approval under Austrian law. The present study analyzes updated versions of the same datasets, together with publicly available wastewater and aggregated mobility data.

