## Supplementary Information for "Wastewater sequence data reveal national and cross-border virus transmission dynamics"

### A Robustness analyses in simulations

#### A.1 The mutation-prevalence weight exponent $\delta$

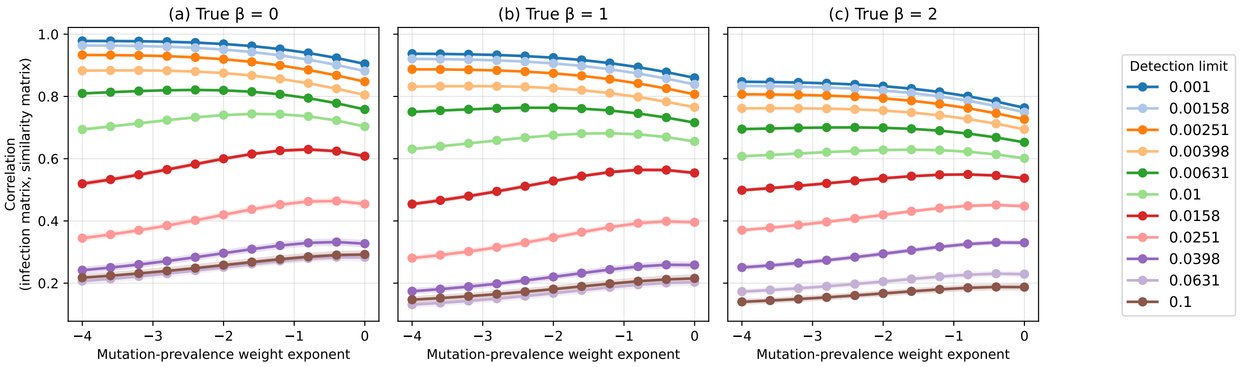


Figure A.1: **Sensitivity of CAP counting to the mutation-prevalence weighting exponent.** Correlation between the true transmission matrix and the inferred CAP counting matrix as the mutation-prevalence weight exponent $\delta$ is varied for simulations with true distance-decay exponents (a) $\beta_{\text{dist}} = 0$, (b) $\beta_{\text{dist}} = 1$ and (c) $\beta_{\text{dist}} = 2$. Colors indicate the allele-frequency detection limit. Points and lines show means across 500 independently seeded simulations, and shaded regions show 95% normal-approximation confidence intervals. The inverse-prevalence weighting used in the main analysis is defined in Eq. 5.

#### A.2 Sensitivity and perturbation analyses

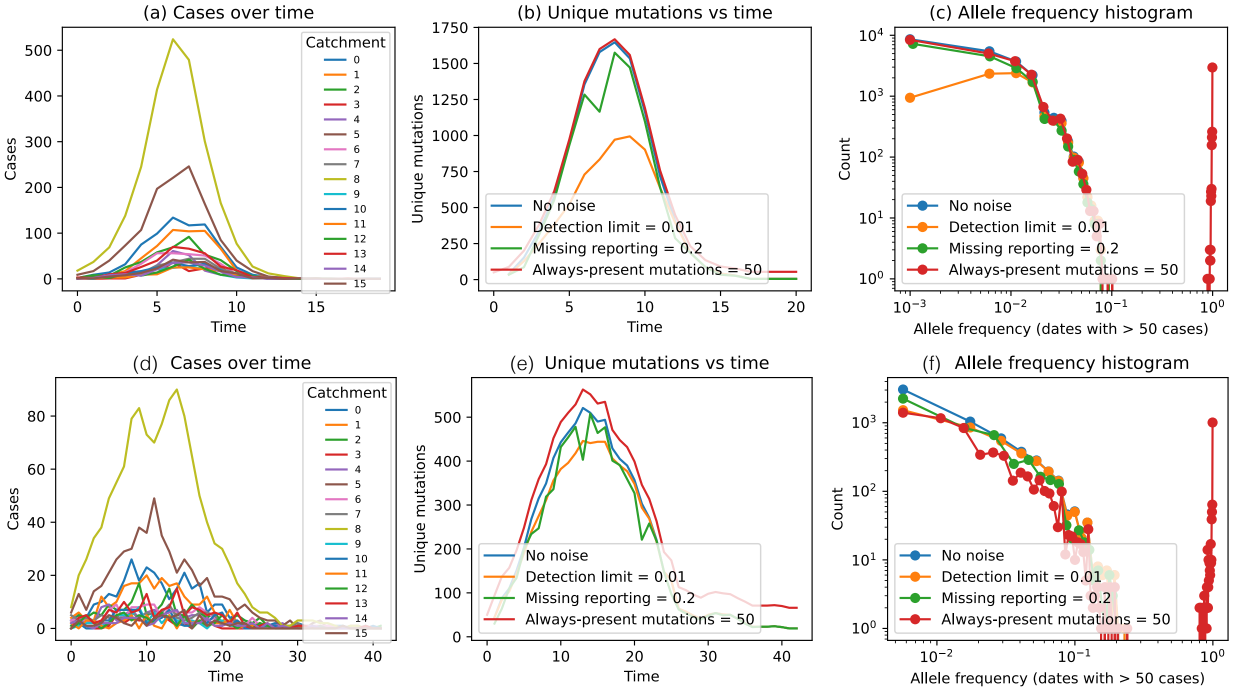


Figure A.2: **Effects of synthetic observation noise.** One representative synthetic epidemic showing (a) catchment-level case counts over time, (b) the number of observed unique mutations over time and (c) allele-frequency distributions on dates with more than 50 cases. In (b) and (c), blue denotes complete observation, orange a detection limit of 0.01, green 20% missing catchment–time reports and red 50 apparently fixed mutations. (d)-(f) mirrors (a)-(c) except with $R_0 = 1.25$ instead of the default value $R_0 = 2.0$. The comparison of the orange curves in panels (b) and (c) demonstrate how a larger number of infections can hide individual mutations at lower frequencies below the allele-frequency detection limit. The observation model is described in Methods 1.3.

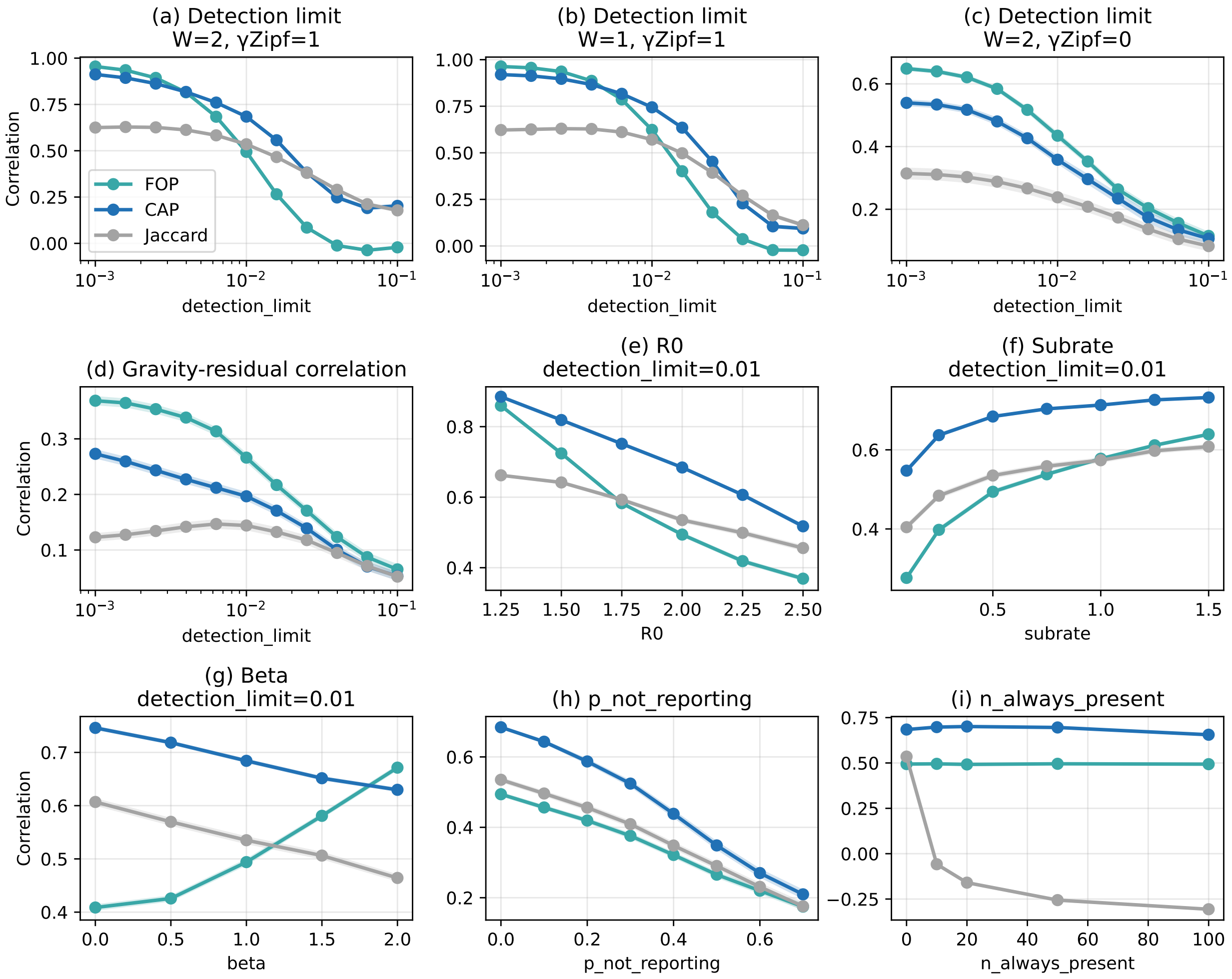


Figure A.3: **Correlation between the true transmission matrix and ETMs inferred by FOP counting (teal), CAP counting (blue) and temporally resolved Jaccard similarity (grey).** (a) Correlation across allele-frequency detection limits under the baseline wastewater observation model as in Fig 2c. (b) The same analysis using an observation model in which an infected individual's sequence contributes to the synthetic wastewater observation only at the time of infection ($W = 1$). (c) Performance across detection limits for equal population sizes ($\gamma_Z = 0$, $W = 2$). (d) Correlation between gravity-adjusted residuals of the inferred ETMs and the true transmission matrix across allele-frequency detection limits. Residuals were obtained by fitting the gravity model separately to each matrix and subtracting the population- and distance-associated component from log-transformed edge weights; higher values therefore indicate better recovery of edge-level deviations from the expected gravity structure. (e–i) Performance as (e) the basic reproduction number $R_0$, (f) the expected substitutions per transmission $\mu$, (g) the true distance-decay exponent $\beta_{\text{dist}}$, (h) the missing-reporting probability and (i) the number of apparently fixed mutations are varied. Points and lines show means across independently seeded simulations, and shaded regions show 95% normal-approximation confidence intervals. The main analyses used 500 simulations; the equal-population analysis in (c) used 100 simulations. Unspecified parameters were held at the baseline values given in Methods 1.3.

### B Recovery of the Distance-Decay Exponent

The distance-decay exponent inferred from an ETM is attenuated relative to the exponent imposed on the synthetic contact matrix for two reasons (Supplementary Fig. B.1). First, the inferred ETMs scale sublinearly with the realized transmission matrix, with ETM $\propto$ TM$^{\gamma}$ and $\gamma < 1$ across the simulated range. This compression is stronger at the higher allele-frequency detection limit and differs among ETM construction methods. Second, even when the gravity model is fitted directly to the realized transmission matrix, the inferred distance-decay exponent is smaller than the exponent imposed on the underlying contact matrix. Consequently, attenuation arises both from the mapping between the contact matrix and realized transmission and from reconstruction of transmission structure from mutation observations. Lowering the allele-frequency detection limit reduces, but does not eliminate, the latter effect.

We therefore interpret the empirical quantity $\beta_{\text{eff}}$ as a measure of relative spatial transmission structure rather than as an unbiased estimate of the distance-decay exponent of the underlying contact process.

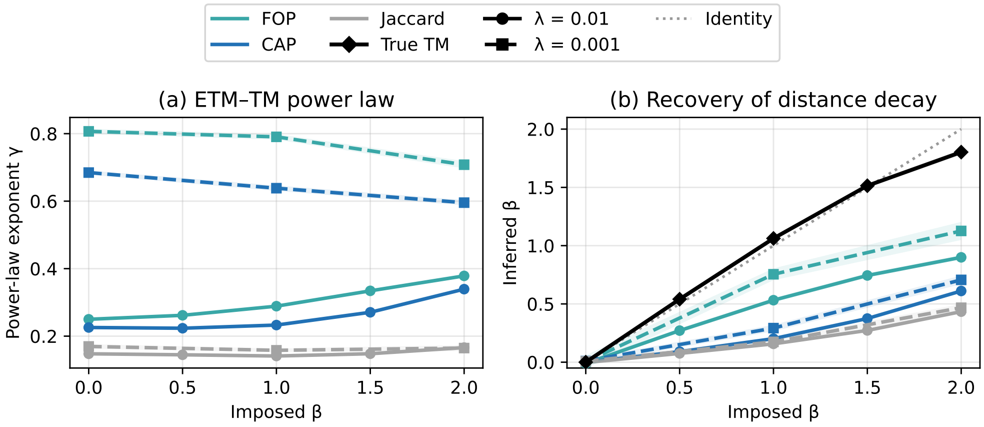


Figure B.1: **Recovery of the distance-decay exponent from synthetic ETMs.** (a) Power-law exponent $\gamma$ obtained from ETM $= c\,\text{TM}^{\gamma}$ for FOP counting (teal), CAP counting (blue), and temporally resolved Jaccard similarity (grey) as the distance-decay exponent imposed on the synthetic contact matrix is varied. Values below one indicate compression of variation in the realized transmission matrix by the inferred ETM. (b) Distance-decay exponent obtained by fitting the gravity model either directly to the true realized transmission matrix (black) or to the inferred ETMs. The dotted line denotes identity. Solid lines with circles show results for an allele-frequency detection limit of $\lambda = 0.01$, whereas dashed lines with squares show results for $\lambda = 0.001$. The true transmission matrix is independent of the observation detection limit and is therefore shown once. Lines and points show means across 50 independently seeded simulations and shaded regions show 95% $t$-based confidence intervals.

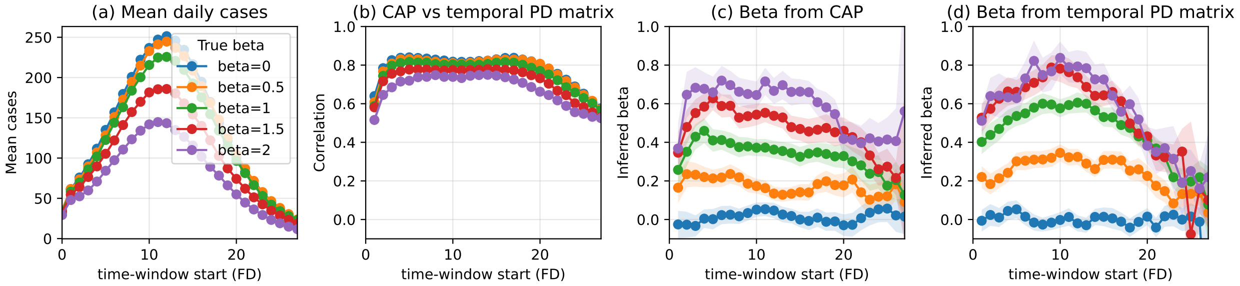


Figure B.2: **Temporal validation of CAP counting in synthetic epidemics with $R_0 = 1.25$.** (a) Mean daily case incidence across the simulated epidemic. (b) Correlation between the CAP counting matrix and the true window-specific transmission matrix. Distance-decay exponents fitted to (c) the CAP counting matrix and (d) the true parent–destination matrix. Colors indicate the imposed distance-decay exponent. Estimates were calculated in three-time-step windows; points and lines show means across 100 independently seeded simulations, and shaded regions show 95% normal-approximation confidence intervals.

### C Detection of reductions in transmission

#### C.1 Model implementation details

For both the synthetic and Austrian applications, location effects were parameterized using the zero-sum non-centred parameterization

$$z_i \sim \mathcal{N}(0,1), \qquad u_i = \tau \left( z_i - \frac{1}{K} \sum_{k=1}^{K} z_k \right),$$

so that $\sum_i u_i = 0$. We used $\alpha \sim \mathcal{N}(\overline{\ell(S)}, 2^2)$, $\beta \sim \mathcal{N}(0,1)$, $\tau \sim \text{HalfNormal}(0.5)$ and $\sigma \sim \text{HalfNormal}(0.5)$, where $\overline{\ell(S)}$ denotes the mean of $\log(S_{ij})$ over the positive, finite, unordered location pairs included in the model. In the synthetic analysis, independent $\mathcal{N}(0,1)$ priors were used for the population and distance coefficients. Posterior sampling used four chains with 1,000 tuning iterations and 1,000 retained draws per chain and a target acceptance probability of 0.92.

For the synthetic perturbation analysis, FOP counting and true realized transmission matrices were symmetrized as $(M + M^{\mathsf{T}})/2$ before fitting, whereas CAP counting was already symmetric. Only positive finite unordered pairs were included. For each replicate, we recorded the posterior probability $P(u_i < 0 \mid D)$ for the truly perturbed location. Detection thresholds were calibrated separately for each analysis setting as the 95th percentile of this probability in unperturbed simulations. Detection probabilities were calculated across simulation replicates, with 95% Wilson binomial confidence intervals.

#### C.2 Synthetic results

To test whether ETMs could resolve local changes in transmission, we imposed progressively stronger reductions in transmission involving a single subpopulation and evaluated detection of a negative location effect at the perturbed site. Detection thresholds were calibrated from null simulations to an approximately 5% false-detection rate. Detection probability increased with perturbation magnitude (Supplementary Fig. C.1). Moderate reductions were detected with high probability when the true realized transmission matrix was observed directly, whereas ETM-based inference generally required larger perturbations, particularly at the finite allele-frequency detection limit $\lambda = 0.01$. For sufficiently strong reductions, both FOP counting and CAP counting achieved high detection probability.

The lower sensitivity to local perturbations contrasted with the high overall correspondence between inferred and true transmission matrices. After removing the population- and distance-associated gravity component, correlations between the residual edge structure of inferred and true transmission matrices were markedly lower and decreased further as the allele-frequency detection limit increased (Supplementary Fig. A.3d). Thus, high recovery of global transmission structure did not imply equivalent recovery of local deviations from that structure.

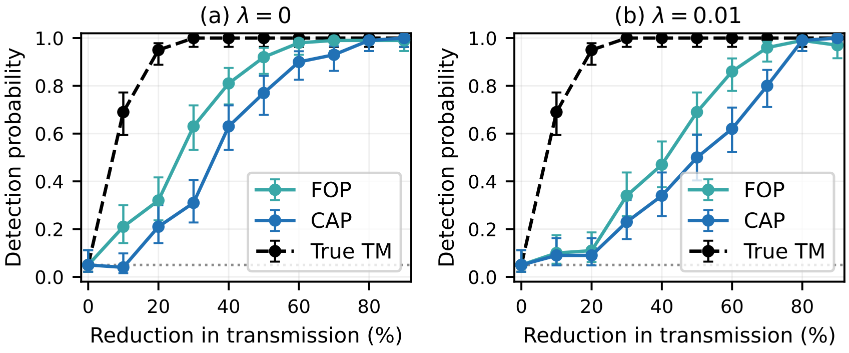


Figure C.1: **Detection of reductions in transmission.** Detection probability for the perturbed location as a function of the imposed reduction in transmission involving that location, under complete mutation observation ($\lambda = 0$; a) and a finite allele-frequency detection limit ($\lambda = 0.01$; b). FOP counting and CAP counting are compared with inference from the true realized transmission matrix (True TM). Detection probabilities were estimated from 100 independently seeded simulations per perturbation level; error bars show 95% Wilson binomial confidence intervals.

### D Viral reference values for $R_e$ and $\mu$

Table D.1: **Viral reference coordinates used in Fig. 2d.** $R_e$ values are context-specific estimates for the indicated epidemic, outbreak or season. Values in parentheses give reported uncertainty intervals or ranges where available. For H1N1, the plotted value is the midpoint of the reported early-$R_t$ range. For Zika, an outbreak-onset estimate of $R_0$ is shown. The parameter $\mu$ is the expected number of substitutions per genome per transmission used for plotting.

| Virus | Setting / period | Estimate type | $R_e$ [ref.] | $\mu$ [ref.] |
|---|---|---|---|---|
| H1N1 (2009) | Hong Kong, 2009 | early $R_t$ | 1.45 (midpoint of 1.4–1.5) [1] | 0.32 [2] |
| H3N2 (2011–12) | Italy, 2011–2012 | seasonal $R_e$ | 1.31 (1.27–1.34) [3] | 0.15 [2] |
| RSV (2018–19) | Germany, 2018–2019 | seasonal $R_e$ | 1.99 [4] | 0.20 [2] |
| Ebola (2014) | Sierra Leone, 2014 | phylodynamic $R_e$ | 1.26 (1.04–1.54) [5] | 0.72 [2] |
| Zika (2015) | Salvador, Brazil, 2015 | outbreak-onset $R_0$ | 2.10 (1.8–2.5) [6] | 0.63 [2] |
| Norovirus (2009–17) | United States, 2009–2017 | median outbreak $R_e$ | 1.29 (IQR 1.12–1.74) [7] | 0.39 [8, 9] |
| SARS-CoV-2 (2020) | Wuhan, January 2020 | early $R_t$ | 2.35 (1.15–4.77) [10] | 0.52 [2] |
| Measles (1996) | Wincrange, Luxembourg, 1996 | early-outbreak $R_e$ | 1.50 (0.9–2.2) [11] | 0.26 [2] |
| Mumps (2016) | Harvard University, 2016 | outbreak $R_e$ | 1.70 (1.50–1.91) [12] | 0.65 [2] |
| Delta (2021) | Austria, 19 July 2021 | variant-specific $R_e$ | 1.34 (1.29–1.39) [13] | 0.52 [2] |
| Omicron (2021) | Austria, 20 December 2021 | variant-specific $R_e$ | 1.71 (1.62–1.81) [14] | 0.44 [2] |

### E Results on US clinical sequences and SafeGraph data

#### GISAID EpiCoV genetic database

We downloaded the entire GISAID EpiCoV database for the United States between March 2020 and March 2023 [15]. Our analysis mainly relies on the amino-acid-level substitution dataset of each sequence compared to the WIV04 reference sequence collected in late 2019 in Wuhan. Although the amino-acid-level substitution data is more aggregated than the raw genetic data (three nucleotides encode one amino acid, with multiple triplets having the same encoding), it still contains highly detailed information about the genetic code of the samples. We filtered out samples where the substitution data was not computed on the full length virus genome. Also, the state of Idaho was excluded from the analysis because its mean number of amino-acid substitutions was markedly higher than that of other states at comparable collection dates.

For comparison with mobility, we downloaded monthly state-to-state SafeGraph population-flow matrices [16, 17] and fitted the same gravity-model distance exponent used for the ETMs. Within-state flows, Puerto Rico and Idaho were excluded. We compared the resulting SafeGraph exponent with the ETM-derived $\beta_{\mathrm{eff}}$, international flights, Google mobility, case counts and mutation counts using monthly Pearson correlations.

The United States analysis showed a temporal pattern in $\beta_{\mathrm{eff}}$ that was positively associated with the SafeGraph distance exponent and negatively associated with international flights (Supplementary Fig. E.1). Since these data were clinical sequences aggregated to emulate wastewater observations, this analysis is presented as an illustrative extension of the wastewater results.

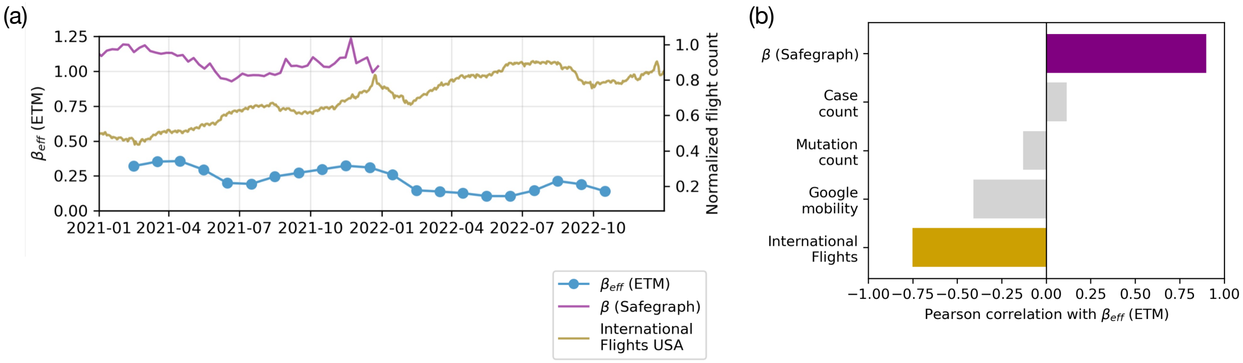


Figure E.1: **Temporal ETM structure in United States clinical sequences and comparison with mobility.** (a) Effective distance-decay exponent $\beta_{\mathrm{eff}}$ estimated from ETMs in rolling three-month windows (blue), the corresponding SafeGraph mobility-derived distance exponent (purple) and normalized international flight counts (gold). (b) Pearson correlations between monthly $\beta_{\mathrm{eff}}$ estimates and the SafeGraph exponent, case count, mutation count, Google mobility and international flights. The SafeGraph and flight correlations are highlighted in purple and gold, respectively; the remaining covariates are shown in grey. Idaho was excluded from the analysis. Exact correlation statistics are provided in Supplementary Table E.1.

Table E.1: **Correlations between spatial transmission localization and mobility and epidemiological covariates in the United States.** Pearson correlation coefficients ($r$) between monthly effective distance-decay estimates ($\beta_{\mathrm{eff}}$) and the indicated covariates, corresponding to Supplementary Fig. E.1. Correlations were calculated using pairwise-complete monthly observations. Degrees of freedom are $n-2$. All $P$ values are two-sided. No correction for multiple comparisons was applied.

| Variable | $r$ | $n$ | df | $P$ value |
|---|---|---|---|---|
| International flights | −0.766 | 23 | 21 | 0.000021 |
| Google mobility | −0.406 | 21 | 19 | 0.067995 |
| Mutation count | −0.222 | 23 | 21 | 0.309317 |
| Case count | 0.073 | 23 | 21 | 0.742335 |
| $\beta$ (SafeGraph) | 0.884 | 11 | 9 | 0.000304 |

### F Additional figures for real data

#### F.1 Results for the Austrian wastewater dataset

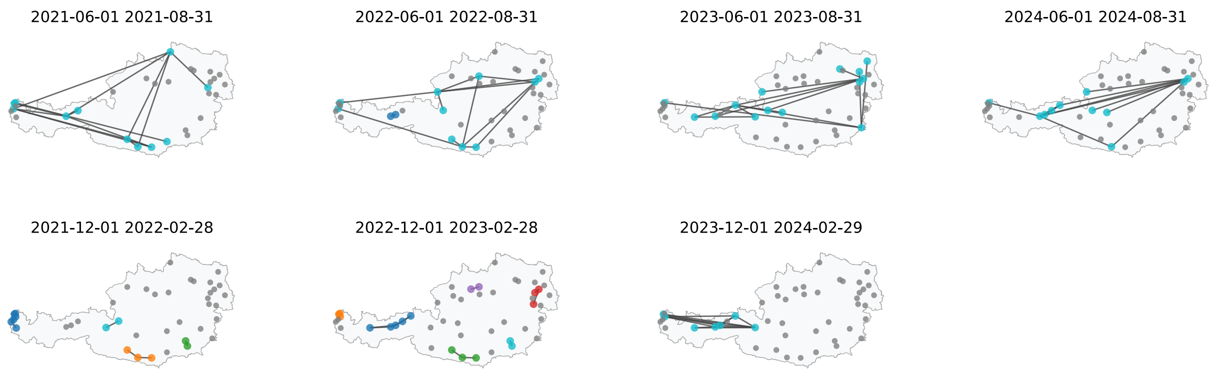


Figure F.1: **Seasonal Austrian ETM networks.** Networks of the 15 strongest positive ETM connections for four summer and three winter periods. Node colors identify connected components of the displayed links; smaller grey nodes denote reporting catchments not incident to one of the selected connections. The three networks used in Fig. 3a–c are included in this series.

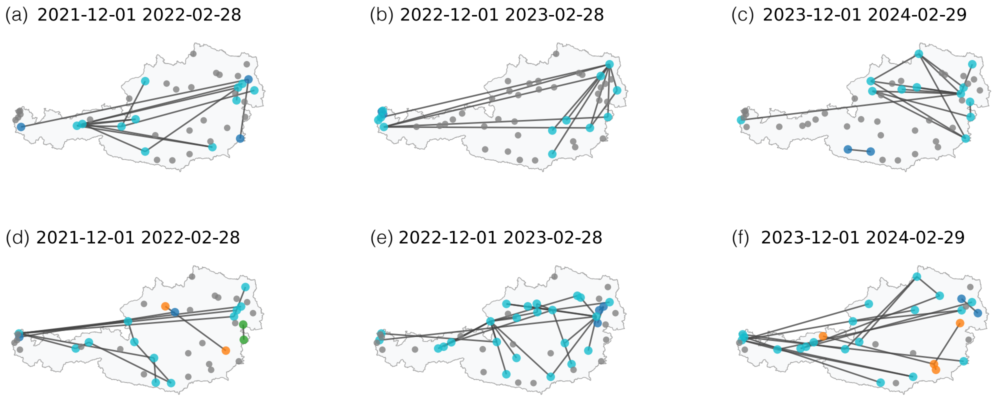


Figure F.2: **Seasonal Austrian ETM networks with Jaccard and FOP counting.** Networks of the 15 strongest positive ETM connections three winter periods computed with Jaccard similarity in panels (a)-(c) and FOP counting in panels (d)-(f).

Table F.1: **Correlations between spatial transmission localization and mobility, sequencing, and epidemiological covariates.** Pearson correlation coefficients ($r$) between monthly effective distance-decay estimates ($\beta_{\text{eff}}$) and the indicated covariates, corresponding to Fig. 3e. Correlations were calculated using pairwise-complete monthly observations. Degrees of freedom are $n-2$. All $P$ values are two-sided. No correction for multiple comparisons was applied.

| Variable | $r$ | $n$ | df | $P$ value |
|---|---|---|---|---|
| Google mobility | −0.712 | 21 | 19 | 0.000297 |
| Meta mobility | −0.581 | 23 | 21 | 0.003634 |
| Overnight stays | −0.339 | 45 | 43 | 0.022692 |
| Flights | −0.227 | 34 | 32 | 0.197363 |
| Mutation count | 0.228 | 45 | 43 | 0.131734 |
| WW viral load | 0.232 | 45 | 43 | 0.124498 |

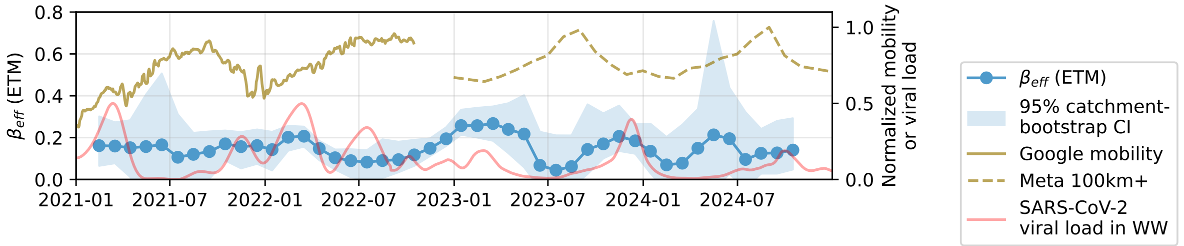


Figure F.3: **Temporal Austrian distance-decay estimates with uncertainty.** Effective gravity-model distance exponent $\beta_{\text{eff}}$ in rolling three-month windows, with 95% catchment-bootstrap confidence intervals from 1,000 resamples. Normalized Google mobility, Meta 100-km-plus mobility and SARS-CoV-2 wastewater viral load are shown for comparison.

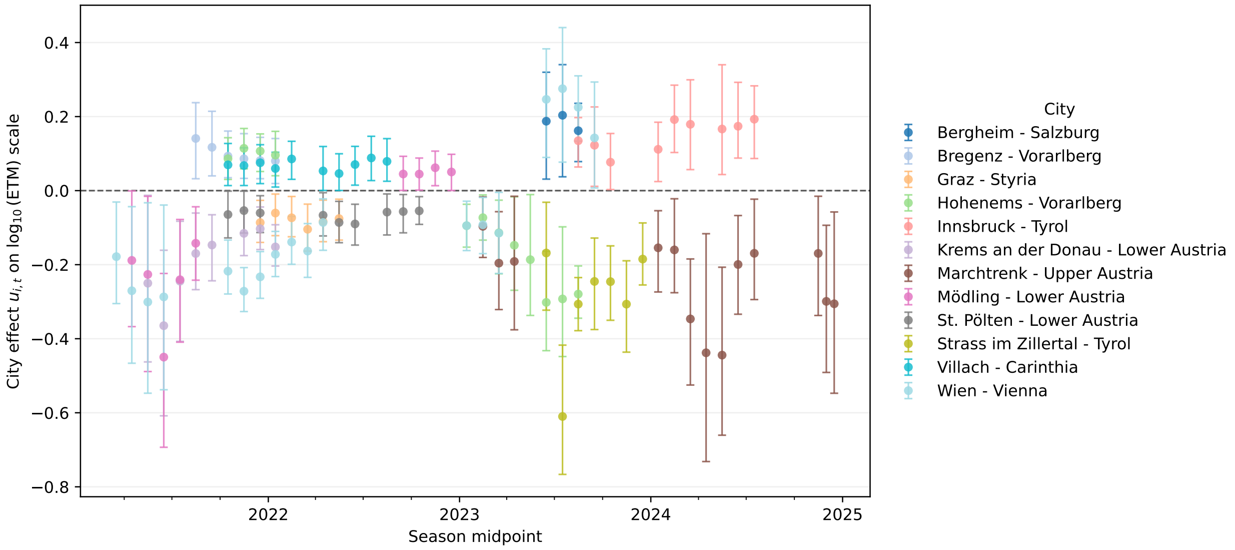


Figure F.4: **Temporally varying commuting-adjusted location effects in Austrian ETMs.** Posterior mean location effects from the hierarchical Bayesian model fitted separately to each seasonal window after adjustment for commuter counts (Supplementary Section C.1). Positive values indicate stronger and negative values weaker ETM connectivity than expected from commuting. Error bars show 99.99% credible intervals, focusing on the strongest patterns in the dataset. The dashed horizontal line at zero denotes no location-specific effect. Only effects whose credible intervals excluded zero with the same sign for at least three consecutive windows are shown.

#### 78 F.2 Austrian commuting vs ETM comparisons

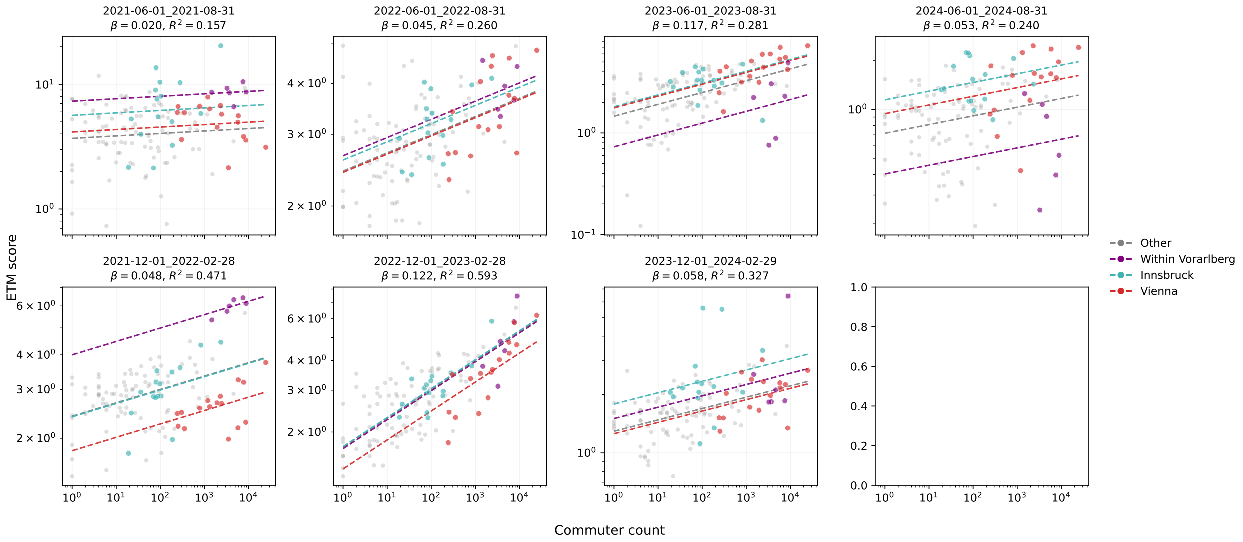


Figure F.5: **Seasonal relationship between Austrian ETMs and commuting.** Positive ETM values versus commuter counts for seven selected summer and winter periods. Colors distinguish pairs involving Vienna, pairs involving Innsbruck, within-Vorarlberg pairs and all remaining pairs; dashed lines show fits of Eq. 16. The same 126 unordered catchment pairs were used in every displayed period.

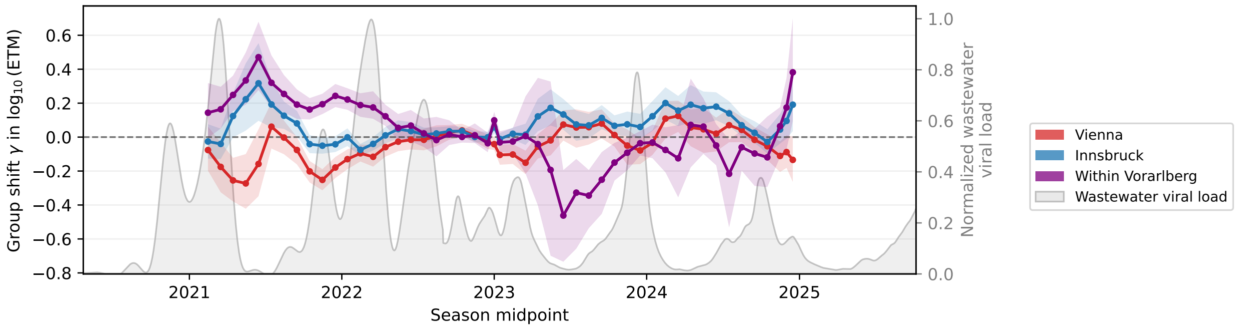


Figure F.6: **Temporal commuting-adjusted ETM shifts in Austria.** Group-specific shifts $\gamma$ for Vienna, Innsbruck and within-Vorarlberg pairs estimated in rolling windows using Eq. 16. Shaded regions show 95% pair-bootstrap confidence intervals from 2,000 resamples, and the grey background shows normalized SARS-CoV-2 wastewater viral load. The analysis used the 106 unordered catchment pairs available in all 49 windows.

#### F.3 Austrian-Swiss cross-border dataset

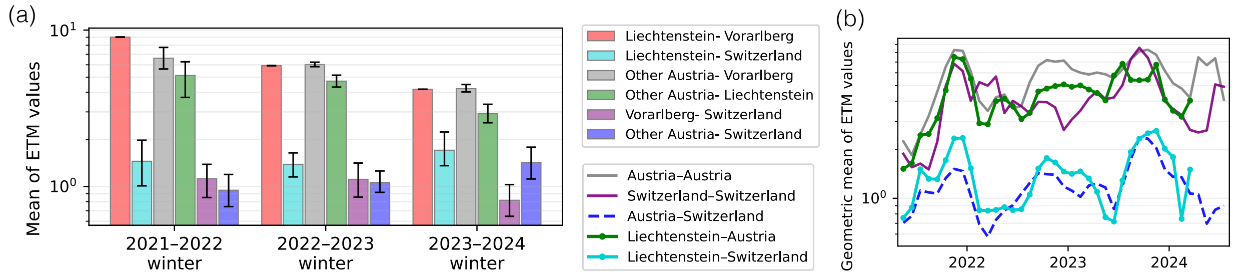


Figure F.7: **Sensitivity analysis of cross-border ETM connectivity using a balanced subset of Austrian and Swiss catchments.** (a) Geometric mean of positive ETM values for the same regional pair classes shown in Fig. 4e, after restricting the analysis to a balanced set of well-sampled Austrian and Swiss catchments. Error bars show 95% catchment-bootstrap confidence intervals from 2,000 resamples. The elevated connectivity of Liechtenstein with Austrian catchments, including Vorarlberg, persists after balancing sampling coverage, indicating that unequal sampling density alone does not explain the Liechtenstein–Austria pattern. (b) Geometric mean of positive ETM values in rolling three-month windows for within-Austria, within-Switzerland, Austria–Switzerland, Liechtenstein–Austria and Liechtenstein–Switzerland catchment pairs in the balanced dataset. Connections crossing between the Austrian/Liechtenstein and Swiss datasets show persistently lower ETM values than same-dataset comparisons.

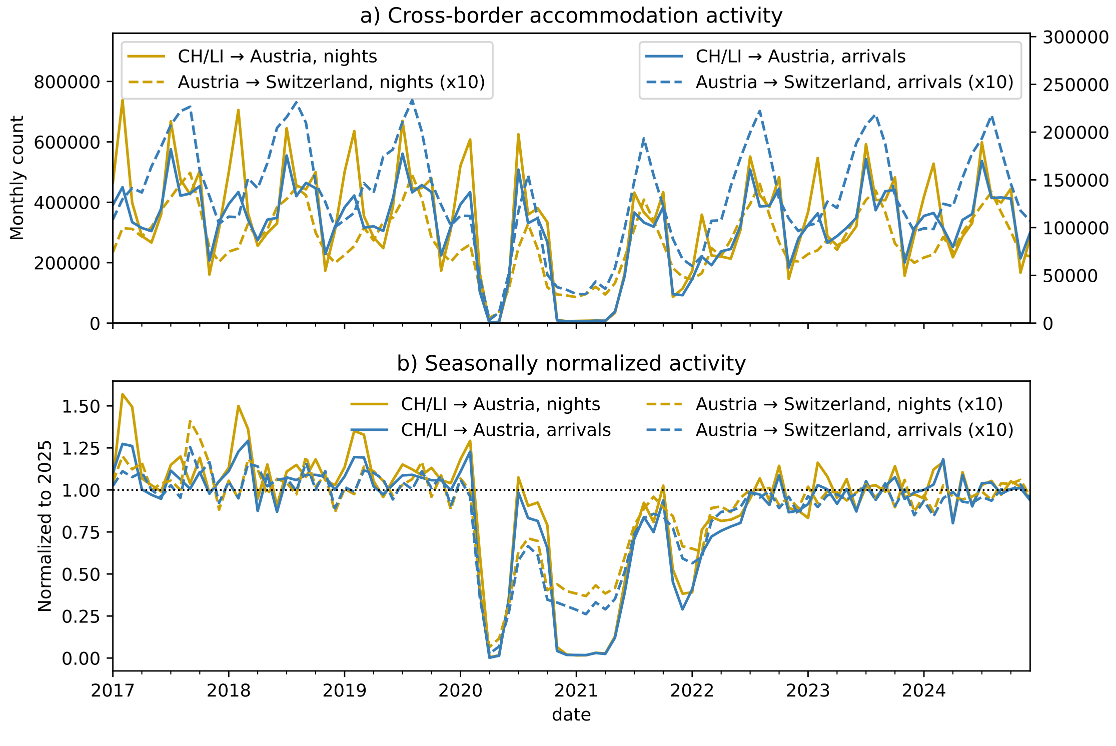


Figure F.8: Cross-border accommodation activity between Austria and Switzerland. (a) Monthly overnight stays and arrivals for visitors from Switzerland and Liechtenstein travelling to Austria and for Austrian residents travelling to Switzerland. Overnight stays are shown in gold and arrivals in blue. Solid lines indicate travel to Austria and dashed lines travel to Switzerland. For visual comparison, the Austria-to-Switzerland series were multiplied by 10 in this panel only. Absolute levels should not be compared directly between directions because the Austrian and Swiss sources do not cover identical accommodation sectors. (b) The same series after calendar-month normalization to 2025. Each observation was divided by the value recorded in the corresponding month of 2025, such that 1 denotes the 2025 monthly level. A post-pandemic reference year was used because the raw series show that the seasonal pattern observed in 2017–2019 differs from that in later unrestricted years, and normalization to the pre-pandemic period would therefore retain residual seasonality. The analysis period ends in December 2024, making 2025 an out-of-sample reference year.

#### G Analytic results for FOP counting

Consider a sequence of epidemics indexed by population size $N \to \infty$ in a metapopulation with catchments $\{1, \ldots, K\}$. Fix a time $t$. Assume:

(A1) **Transmission model.** The epidemic follows a discrete-time metapopulation SIR process in which each infected individual in catchment $i$ is infectious for exactly one time step and then recovers. Conditional on the infected sets, each infected individual in $i$ generates independently, for each $j \neq i$, a transmission to one individual in catchment $j$ at time $t+1$ with probability $p_{ij}$. Let $T_{ij}(t)$ denote the resulting number of transmissions from $i$ to $j$ during step $t \to t+1$.

(A2) **Mutation model.** Each transmission event independently creates a unique mutation with probability $\mu \in (0, 1]$, which is present in the newly infected individual at time $t+1$, and in all of its descendants. The same mutation never arises independently in different lineages (no homoplasy).

(A3) **Perfect detection.** If a mutation is present in infections contributing to wastewater in a catchment at time $t$, it is detected with probability 1.

(A4) **No independent co-occurrence.** The same mutation does not arise independently in two individuals, and there are no additional transmission routes unobserved by the system, e.g. routes through other countries or unobserved catchments.

**Definition 1.** For each ordered pair $i \neq j$, let $\widehat{T}_{ij}(t)$ be the number of mutations first observed anywhere in the metapopulation at time $t$, exclusively in catchment $i$, and subsequently observed in catchment $j$ at time $t+1$.

**Remark G.1.** Under (A2)–(A4), the mutations first observed in catchment $i$ at time $t$ are independent Bernoulli marks, with probability $\mu$, of the individuals infected there at time $t$. By (A1), the $T_{ij}(t)$ transmissions to a fixed catchment $j$ arise from distinct such individuals, and hence

$$\widehat{T}_{ij}(t) \mid T_{ij}(t) \sim \text{Binomial}\big(T_{ij}(t), \mu\big).$$

**Proposition G.1.** Let

$$X = \text{vec}\big(T(t)\big), \qquad Y = \text{vec}\big(\widehat{T}(t)\big),$$

where vectorization runs over all ordered pairs $i \neq j$, and let

$$\bar{X} = \frac{1}{K(K-1)} \sum_i X_i, \qquad \bar{Y} = \frac{1}{K(K-1)} \sum_i Y_i.$$

If, as $N \to \infty$, the empirical variance-to-mean ratio of the off-diagonal transmission counts diverges, i.e.,

$$\frac{\sum_i \big(X_i - \bar{X}\big)^2}{\sum_i X_i} \xrightarrow{p} \infty, \tag{1}$$

then the sample Pearson correlation satisfies

$$\text{corr}(X, Y) = \frac{\sum_i (X_i - \bar{X})(Y_i - \bar{Y})}{\sqrt{\sum_i (X_i - \bar{X})^2}\sqrt{\sum_i (Y_i - \bar{Y})^2}} \xrightarrow{p} 1.$$

*Proof sketch.* Write $Y = \mu X + \varepsilon$, and let $\widetilde{X}$, $\widetilde{Y}$, and $\widetilde{\varepsilon}$ denote the corresponding centered vectors (the realized sample mean of $\varepsilon$ does not need to be zero). Since $\mathbb{E}[\varepsilon \mid X] = 0$ and $\text{Var}(\varepsilon_i \mid X) = \mu(1-\mu)X_i$, the condition Eq. 1 gives

$$\frac{\mathbb{E}[\|\varepsilon\|_2^2 \mid X]}{\mu^2 \|\widetilde{X}\|_2^2} = \frac{(1-\mu)\sum_i X_i}{\mu \sum_i (X_i - \bar{X})^2} \xrightarrow{p} 0.$$

By conditional Markov's inequality and since centering cannot increase the Euclidean norm,

$$\frac{\|\tilde{\varepsilon}\|_2}{\mu \|\tilde{X}\|_2} \xrightarrow{p} 0.$$

Using $\widetilde{Y} = \mu \widetilde{X} + \widetilde{\varepsilon}$, Cauchy–Schwarz and the reverse triangle inequality give, respectively,

$$\left| \frac{\langle \widetilde{X}, \widetilde{Y} \rangle}{\|\widetilde{X}\|_2^2} - \mu \right| = \frac{|\langle \widetilde{X}, \widetilde{\varepsilon} \rangle|}{\|\widetilde{X}\|_2^2} \leq \frac{\|\widetilde{\varepsilon}\|_2}{\|\widetilde{X}\|_2} \xrightarrow{p} 0, \quad \text{and} \quad \left| \frac{\|\widetilde{Y}\|_2}{\|\widetilde{X}\|_2} - \mu \right| = \frac{\big|\|\widetilde{Y}\|_2 - \|\mu \widetilde{X}\|_2\big|}{\|\widetilde{X}\|_2} \leq \frac{\|\widetilde{Y} - \mu \widetilde{X}\|_2}{\|\widetilde{X}\|_2} \xrightarrow{p} 0.$$

Finally, since $\mu > 0$,

$$\mathrm{corr}(X, Y) = \frac{\langle \widetilde{X}, \widetilde{Y} \rangle}{\|\widetilde{X}\|_2 \, \|\widetilde{Y}\|_2} = \frac{\langle \widetilde{X}, \widetilde{Y} \rangle / \|\widetilde{X}\|_2^2}{\sqrt{\|\widetilde{Y}\|_2^2 / \|\widetilde{X}\|_2^2}} \xrightarrow{p} \frac{\mu}{\sqrt{\mu^2}} = 1.$$

□

**Remark G.2** (Interpretation of the proposition)**.** In this idealized model, the mutation count $\widehat{T}_{ij}(t)$ is obtained by independently Bernoulli-thinning the true transmission count $T_{ij}(t)$ with probability $\mu$. The proposition says that if the variance across true transmission counts grows faster than their mean, then the inferred ETM becomes asymptotically perfectly correlated with the true transmission matrix.

**Remark G.3** (Interpretation of the condition)**.** The variance-to-mean divergence condition is mild in heterogeneous metapopulation settings. It is naturally satisfied when transmission counts scale with population size and exhibit persistent differences in their expected intensities, as may arise from heterogeneous catchment population sizes, infection prevalence, or transmission probabilities.

**Remark G.4** (Multiple offspring)**.** The assumption in (A1) can be relaxed to allow multiple offspring per transmitting parent and destination. For example, suppose the positive offspring numbers of transmitting parents are independent across parents, independent of mutation marks, and have a common distribution across all catchment pairs, with mean $r$ and finite variance. Then, while FOP counting counts mutation-marked transmitting parents, the preceding second-moment argument extends to individual transmissions with proportionality factor $\mu/r$, giving the same correlation conclusion.
